# Interoceptive mechanisms of body image disturbance in anorexia nervosa: Study protocol of a randomized parallel-group trial

**DOI:** 10.64898/2026.10.26.26366139

**Authors:** E. A. Bernardini, E. M. Choquette

## Abstract

**Background:** Current treatments for anorexia nervosa (AN), a dangerous psychiatric disorder, show only moderate efficacy. Interoceptive processing dysfunctions (i.e., how the brain receives and processes body afferents), and body image disturbance (BID) are clinical features of AN. Floatation-REST (Reduced Environmental Stimulation Therapy) has been reported to enhance both subjective interoceptive experiences and BID. The proposed project works to establish (1) how interoceptive processing dysfunctions may contribute to BID and (2) how the combination of Floatation-REST with psychotherapeutic principles may enhance interoceptive processing and BID in AN.

**Methods:** In this randomized parallel-group (NCT06332963), single-site trial, 102 women and girls with AN will be recruited from an inpatient program. After completing baseline measurements, participants are randomized on a 1:1 ratio to receive either the interoceptively focused treatment (IFT) or exteroceptively focused treatment (EFT) condition. Both conditions utilize acceptance- and mindfulness-based principles, however incorporate skills that focus on interoception or exteroception, respectively. Participants complete acute measures following each experimental therapeutic session, as well as post intervention, six-week and six-month follow-up. The primary outcome of the current study is acute changes in BID assessed by the Photographic Figure Rating Scale. Additional outcomes include acute changes in interoception, and longitudinal intervention effects on self-reported, behavioral, and neuroimaging measures of AN symptoms, BID, and interoception.

**Discussion:** If interventions demonstrate reductions in BID, mechanisms of BID will be further understood, informing the development of future therapies for BID in AN.

## Introduction

Anorexia nervosa (AN) is a rare, but deadly psychiatric disorder (∼1.4% lifetime prevalence), which accounts for more than 10,000 deaths per year in the United States alone (Harris and Barraclough, 1998; Hoang et al., 2014; Galmiche et al., 2019; Deloitte Access Economics, 2020). Despite the intense delivery of treatments for AN via inpatient settings, as many as 50% of individuals with AN will relapse within the first year of treatment (Khalsa et al., 2017), and 20% will remain chronically ill (Steinhausen, 2002). A core diagnostic feature of AN is body image disturbance (BID; American Psychiatric Association, 2022). BID tends to be highly resistant to change in AN (Chapa et al., 2020), and is integral to the maintenance and relapse of the disorder (Carter et al., 2004, 2012; Keel et al., 2005). Body image is comprised of both cognitive/affective (how one thinks and feels about their body) and perceptual (the mental representation of one’s body size/shape) components (Cash and Green, 1986; Cash and Deagle, 1997; Cash, 2004; Cash and Smolak, 2012). In AN, perceptual BID manifests as an overestimation of one’s body size (Gardner and Brown, 2014; Mölbert et al., 2017). It has been posited that the construction of body image is a process of sensing, interpreting, and incorporating both external and internal bodily signals which integrate with cognitive schemas to create a representation of one’s body (Choquette and Khalsa, 2025b).

Considering this process of body image perception, it is theorized that perceptual BID may be a consequence of abnormal interoceptive processing (i.e., difficulties with sensing and integrating signals), which prompts an overreliance on external signals, such as visual input (Riva et al., 2012; Gadsby, 2017; Stanghellini et al., 2019; Choquette and Khalsa, 2025b). This in turn may result in negative self-objectification (i.e., seeing one’s body from a third-person perspective as an object) and an inaccurate self-view such as overestimating body size (see chapters Choquette and Khalsa, 2025a, 2025b; Fredrickson and Roberts, 1997; Riva and Gaudio, 2012; Gadsby, 2017; Stanghellini et al., 2019). Not only does interoception seem to contribute to BID, but abnormal interoception is increasingly recognized as playing a central role in the behavioral and physiological expression of AN (Choquette and Khalsa, 2025a). Further, in functional neuroimaging studies, those with AN display signal disruptions in areas of the brain that facilitate interoceptive representation, including the anterior and dorsal mid-insula, as well as the dorsal and ventral striatum (Wagner et al., 2008; Kaye et al., 2009; Kerr et al., 2016). Together, these findings suggest a mechanistic relationship between perceptual BID and interoception; however, no studies to date have examined whether manipulating interoception can modulate perceptual BID in a therapeutic manner.

Treatments exist to reduce BID in AN; however they typically fall short at effectively and consistently improving BID in AN (see review Ziser et al., 2018 and meta-analysis Alleva et al., 2015). Existing interventions tend to emphasize cognitive and affective aspects of BID through techniques such as imagery (Cash and Smolak, 2012), exposure (Cash and Smolak, 2012), and cognitive restructuring (Rosen, 1996). Although approaches such as mirror exposure (Delinsky and Wilson, 2006; Griffen et al., 2018) and virtual reality (Preston and Ehrsson, 2014; Porras Garcia et al., 2019) have been proposed as influencing perceptual BID, findings regarding effectiveness are mixed (Garfinkel et al., 1978; Ziser et al., 2018; Riva et al., 2021).

Moreover, traditional interventions for AN typically do not address deficits in interoceptive processing. Given evidence of interoceptive dysfunction in AN, interventions aimed at increasing sensitivity to interoceptive signaling and engagement with the body may be relevant for improving AN symptomatology. Mindfulness-based practices including yoga, meditation, and body scans (see review Gopan et al., 2024), as well as interventions incorporating mindfulness principles (e.g., acceptance and commitment therapy) have been associated with improvements in subjective interoceptive awareness, although evidence for changes in objective accuracy are less consistent (Fischer et al., 2017; Lima-Araujo et al., 2022). Importantly, interoceptive processing may be particularly relevant to BID because internal bodily signals contribute to how the body is experienced and mentally represented. Together, these findings suggest that mindfulness-based approaches may be useful not only for increasing awareness of bodily signals, but for altering how those signals are attended to and incorporated into bodily experience. The present study builds on this possibility by combining mindfulness-based practices with an experimental manipulation of interoception versus exteroception.

The current intervention draws from two evidence-based therapeutic practices: acceptance and commitment therapy (ACT; Hayes et al., 1999) and mindfulness-oriented recovery enhancement (MORE; Garland, 2013). ACT, a third-wave cognitive behavioral therapy, incorporates mindfulness through activities such as body scans and present-moment awareness, and has demonstrated benefits for cognitive/affective BID symptoms in eating disorders (see meta-analysis Linardon et al., 2019). Secondly, MORE, originally developed for opioid use disorder, is rooted in mindfulness and utilizes exercises such as mindful breath, savoring pleasant experiences, and “zooming in” on bodily experiences (Garland, 2024). Additionally, MORE has been associated with increased interoceptive awareness (Roberts et al., 2022). In the present trial, principles from ACT and MORE are adapted to emphasize either an interoceptive or exteroceptive focus. In the interoceptively focused treatment (IFT) condition, these skills are practiced in the context of Floatation-REST (Reduced Environmental Stimulation Therapy), whereas the exteroceptively focused treatment (EFT) condition externally orients mindfulness practices which are delivered through curated videos.

During Floatation-REST, external sensory input is reduced by attenuating visual, auditory, and somatosensory stimulation, thereby increasing salience of internal bodily sensations. Previous studies have indicated heightened subjective interoceptive awareness post-float (Feinstein et al., 2018a; Khalsa et al., 2020; Choquette et al., 2023). Additionally, reductions in perceptual BID have been observed acutely following floatation sessions and at six-month follow-up in participants with AN (Khalsa et al., 2020; Choquette et al., 2023). The safety and feasibility have also been established in AN (Khalsa et al., 2020; Choquette et al., 2023). However, it remains unknown whether pairing Floatation-REST with interoceptively focused therapeutic skills produces greater improvements in BID than an exteroceptively focused mindfulness intervention.

The current trial is designed to examine the role of interoceptive processing in BID, while providing a proof-of-concept evaluation of a novel mindfulness-based treatment developed to target and enhance this relationship. This trial uniquely and systematically examines associations between BID and interoception using multiple levels of analysis (i.e., self-report and behavioral assays and neuroimaging) and combines them with experimental perturbations of interoceptive or exteroceptive processing to examine the unique impact of interoception on perceptual BID. First, we aim to examine how an interoceptive manipulation acutely changes BID in AN. Secondly, we aim to determine cross-sectional multilevel associations between interoception and BID in AN. Finally, exploratory aims include the longitudinal, mechanistic mediators, and neural signatures of BID.

## Methods and Design

Written in accordance with the SPIRIT guidelines (Chan et al., 2025), this protocol describes an ongoing, randomized, parallel-group, exploratory trial of an experimental therapeutic intervention for women and girls with AN. The study is funded by the National Institute of Mental Health (K23 MH135213) and registered on ClinicalTrials.gov (NCT06332963). Research is conducted at a single site at the Laureate Institute for Brain Research (Tulsa, OK), and data is collected by staff who have completed human subject training. Prior to study participation, informed consent is obtained from participants aged 18 years or older, whereas parental or guardian permission and participant assent are obtained for minors aged 13–17 years.

Following an initial screening and consent visit (V0), participants complete eight in-person sessions (V1-V8) and two electronic follow-up sessions (V9 and V10; see Figure 1 for study flow). During baseline sessions, participants complete physiological, behavioral, neuroimaging, and self-report data collection (see Table 1 for full measures by visit).

**Figure 1.**
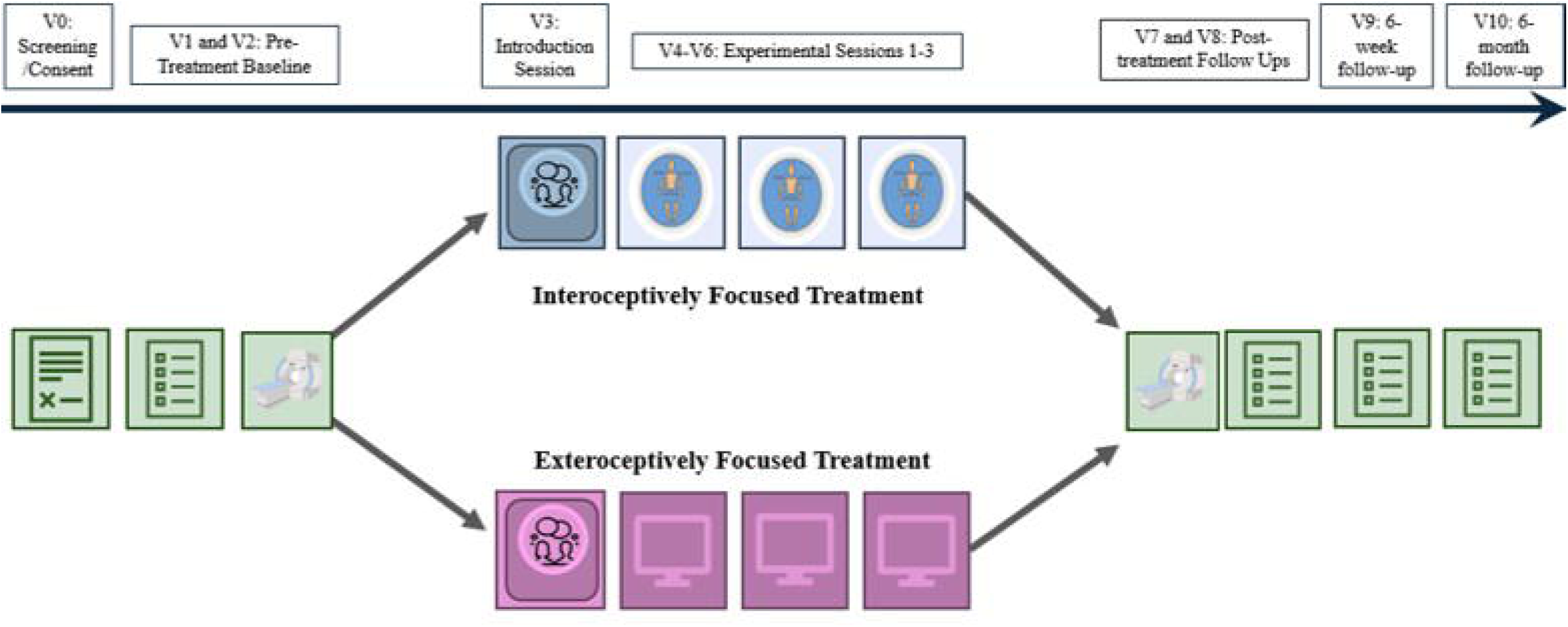
Diagram of Study Protocol. *Note*. Figure represents study flow which includes screening, pre-intervention behavioral and functional magnetic resonance imaging (fMRI) sessions, introduction session, three experimental conditions, post-intervention behavioral and fMRI sessions, as well as 6-week and 6-month follow-ups.

**Table 1.** Study Schedule and Measures.

|  |  | V0: Enrollment | V1 and V2:<br>Pre-Treatment<br>Baseline | V3:<br>Introduction<br>Session | V4-V6:<br>Experimental<br>Sessions 1-3 | V7 and V8:<br>Post-<br>treatment<br>Follow-Ups | V9: 6-Week<br>Follow-Up | V10: 6-Month<br>Follow-Up |
| --- | --- | --- | --- | --- | --- | --- | --- | --- |
| **Screening** |  |  |  |  |  |  |  |  |
| *Instrument* | *Domain* |  |  |  |  |  |  |  |
| Informed Consent | Screening | X |  |  |  |  |  |  |
| Demographic Information | Screening | X |  |  |  |  |  |  |
| Psychiatric History | Screening | X |  |  |  |  |  |  |
| Medical History | Screening | X |  |  |  |  |  |  |
| Urine Screen/Breathalyzer | Screening |  | X |  |  | X |  |  |
| Magnetic Resonance Environment Screen | Screening | X |  |  |  | X |  |  |
| Pre-Treatment Questionnaire | Screening | X |  |  |  |  |  |  |
| C-SSRS (Posner, 2016)* | Screening | X |  |  |  | X |  |  |
| **Intervention** |  |  |  |  |  |  |  |  |
| Introduction Session |  |  |  | X |  |  |  |  |
| IFT or EFT |  |  |  |  | X |  |  |  |
| **Self-Report Assessments** |  |  |  |  |  |  |  |  |
| *Instrument* | *Domain* |  |  |  |  |  |  |  |
| Photographic Figure Rating Scale (PFRS; Swami et al., 2008)† | Perceptual BID | X | X |  | X | X | X | X |
| Eating Disorder Examination Questionnaire (EDE-Q6; Fairburn, 2008) | Eating Disorder Symptom Severity | X | X |  |  | X | X | X |
| Eating Disorder Inventory (EDI; Garner, 1991) | Eating Disorder Symptom Severity |  |  |  |  |  |  | X |
| Body Image State Scale (BISS; Cash et al., 2002) | Cognitive/Affective BID |  | X |  | X | X | X | X |
| Multidimensional Body-Self Relations Questionnaire (MBSRQ; Cash, 2000) | Cognitive/Affective BID |  | X |  |  | X | X | X |
| Multidimensional Assessment of Interoceptive Awareness (MAIA-V2; Mehling et al., 2012) | Interoception |  | X |  |  | X | X | X |
| Interoceptive Attention Scale (IATS; Gabriele et al., 2022) | Interoception |  | X |  |  | X | X | X |
| Visual Analogue Scales (VAS; Feinstein et al., 2018a, 2018b; Khalsa et al., 2020; Choquette et al., 2023) | Interoception (pre- and post-session) |  |  |  | X |  |  |  |
| Somatomap2D Interoception (Ralph-Nearman et al., 2019, 2021; Arevian et al., 2020) | Interoception |  |  |  | X |  |  |  |
| Body Image Acceptance and Action Questionnaire (BI-AAQ-II; Sandoz et al., 2013) | Body Image Flexibility |  | X |  |  | X | X | X |
| State Trait Anxiety Inventory (STAI– State; Spielberger et al., 1970) | Anxiety |  | X |  | X | X | X | X |
| STAI – Trait (Spielberger et al., 1970) | Anxiety |  | X |  |  | X | X | X |
| Anxiety Sensitivity Inventory (ASI-3; Taylor et al., 2007) | Anxiety |  | X |  |  | X |  |  |
| Visceral Sensitivity Index (VSI; Labus et al., 2004) | Anxiety/Gastric Sensitivity |  | X |  |  | X | X | X |
| Illness-Related Words: Anxiety Ratings | Anxiety |  | X |  |  | X |  |  |
| Illness-Related Words: Valence and Arousal | Anxiety/Valence and Arousal |  | X |  |  | X |  |  |
| Patient Health Questionnaire (PHQ-9; Kroenke et al., 2001) | Depression |  | X |  |  | X | X | X |
| Positive and Negative Affect Schedule-Expanded (PANAS-X; Watson et al., 1988) | Affect |  | X |  | X | X | X | X |
| Credibility and Expectancy Questionnaire (CEQ; Devilly and Borkovec, 2000) | Therapy Factors |  | X |  | X<sup>a</sup> |  |  |  |
| Working Alliance Inventory-Self Report (WAI-SR; Horvath and Greenberg, 1989) | Therapy Factors |  |  | X |  | X |  |  |
| Glasgow Sensory Questionnaire (GSQ; Robertson and Simmons, 2013) | Sensory Sensitivity |  | X |  |  | X | X | X |
| ARFID Screener (Zickgraf and Ellis, 2018) | Sensory Sensitivity |  | X |  |  |  |  |  |
| Five Dimension Altered States of Consciousness Scale (5D-ASC; Studerus et al., 2010) | Altered States of Experience |  |  |  | X<sup>a</sup> |  |  |  |
| Body Boundary Dissolution Scale (BBDS; Dambrun, 2016) | Altered States of Experience |  |  |  | X |  |  |  |
| Participant Last Use Summary (PLUS; Lisdahl et al., 2018) | MRI Session (pre-scan) |  | X |  |  | X |  |  |
| Karolinska Sleepiness Scale (KSS; Åkerstedt and Gillberg, 1990) | MRI Session (pre- and post-scan) |  | X |  |  | X |  |  |
| Feedback Interview | Participant Experience |  |  |  |  | X |  | X |
| **Task-Based Assessments** |  |  |  |  |  |  |  |  |
| *Instrument* | *Domain* |  |  |  |  |  |  |  |
| InBody (InBody Co., Ltd., South Korea)/Body Measurements | Physiological Measurements |  | X |  |  | X |  |  |
| Resting State Connectivity | fMRI—Resting State |  | X |  |  | X |  |  |
| Visceral Interoceptive Attention Task (VIA; Simmons et al., 2013) | fMRI—Interoception |  | X |  |  | X |  |  |
| Modified Version of the Adjective Task (Matthews et al., 2010) | fMRI—Self-Referential Processing |  | X |  |  | X |  |  |
| Somatomap3D (Ralph-Nearman et al., 2019, 2021; Arevian et al., 2020) | fMRI—Perceptual BID |  | X |  |  | X |  |  |
| Heartbeat Tapping Task (HBT; Ludwick-Rosenthal and Neufeld, 1985; Canales-Johnson et al., 2015; Smith et al., 2021) | Interoception |  | X |  |  | X |  |  |
| Inspiratory Breath Hold Task (Lapidus et al., 2020) | Interoception |  | X |  |  | X |  |  |
| Somatomap3D (Ralph-Nearman et al., 2019, 2021; Arevian et al., 2020) | Perceptual BID |  | X |  |  | X | X | X |
| Aperture Task (Keizer et al., 2013; Irvine et al., 2019) | Perceptual BID |  | X |  |  | X |  |  |
| String Task (Schneider et al., 2009) | Perceptual BID |  | X |  |  | X |  |  |

*Note.* \*While the C-SSRS is completed at screening/baseline and post-intervention, it may be completed at any other session if there is any indication of suicidal ideation indication (e.g., endorsement of PHQ-9 item 9). †PFRS is the primary study outcome. fMRI = functional magnetic resonance imaging. ‡CEQ completed at baseline and only during the first experimental session, and the 5D-ASC completed only during the second experimental session.

After completion of baseline assessments, participants are randomized on a 1:1 basis to either IFT or EFT. Randomization is stratified by treatment unit (adolescent vs adult; see Supplementary A for further details on randomization). Given the nature of the interventions, participants and therapists cannot be blinded to condition assignment; however, randomization occurs only after the completion of all baseline assessments to reduce potential responding bias.

Following randomization, participants complete a one-hour condition-specific introduction session (V3) followed by three experimental sessions (V4 -V6) according to assigned condition. Intervention procedures are described below. During V4-V6, participants complete pre-and post-session self-report measures. Following experimental sessions, participants complete post-intervention assessments (V7 and V8) which parallel baseline assessments. Finally, participants complete remote follow-up assessments at six weeks (V9) and six months (V10).

### Participants

This study is currently recruiting participants with AN receiving residential care at the Laureate Eating Disorder Program (LEDP; collocated with LIBR). Recruitment is conducted via referrals from LEDP providers and review of the LEDP census. Study staff review the census to identify potentially eligible patients and contact their treatment providers to discuss possible study participation. As of manuscript submission date, we have initially screened 169 individuals and have consented 80 participants. Study eligibility is designed to support participant safety, and inclusion criteria are as follows: (1) primary diagnosis of AN; (2) PFRS (Swami et al., 2008) body dissatisfaction score *≥*1; (3) EDE-Q6 (Fairburn, 2008) Shape Concern Subscale score *≥*3; (4) body mass index **≥**17.5; (5) no current evidence of orthostatic hypotension and/or no evidence of fall risk determined by LEDP provider; (6) residential status of care; (7) no new psychiatric medications in the week prior to randomization; (8) female sex assigned at birth; (9) ages 13-50 years; (10) independently ambulatory; (11) ability to lay flat; (12) English proficiency; (13) willingness/ability to participate in study procedures; (14) provision of informed consent (parent consent and minor assent if less than 18 years of age). Exclusion criteria are as follows: (1) active suicidal ideation with plan and intent; (2) active cutting/skin lacerating behaviors; (3) pregnancy defined by urine screening; (4) acute intoxication (via urine drug screen or breathalyzer); (5) orthostatic hypotension or fall risk determined by provider; (6) seizure reported within the previous 12 months; (7) co-morbid diagnoses of bipolar disorder, schizophrenia, or other psychosis spectrum disorder; (8) systolic blood pressure >160 mmHg; (9) diastolic blood pressure >100 mmHg; (10) resting heart rate < 50 beats per minute. All participants are engaged in ongoing psychotherapeutic and pharmacological (if indicated) care at LEDP. See Supplementary A for further details on recruitment and retention.

The protocol has been revised several times (see Supplementary A Table 1) specifically surrounding eligibility to increase recruitment while prioritizing participant safety. To enhance the ability to recruit participants with enough time to complete the study before discharging, the BMI criteria changed from 18.5 to 17.5. To expand inclusion of potential participants and investigate over a longer age-range, the age range changed from 15-40 to 13-50. Finally, the criteria for orthostatic hypotension now can include an LEDP provider consult to determine if participants are at a clinical fall risk.

### Intervention

Study intervention consists of an introduction session (V3) and three experimental sessions (V4-V6) with a trained therapist. Participants who are randomized to the IFT condition complete three ∼45-minute Floatation-REST sessions paired with interoceptively focused acceptance- and mindfulness-based skills. Participants randomized to the EFT condition complete three ∼45-minute video practice skill sessions focused on present moment and externally focused acceptance- and mindfulness-based skills. Both conditions are delivered individually and in person.

At each experimental session, participants first complete acute self-report measures then receive 15-25 minutes of therapist-guided review of previously learned skills and participants’ experiences practicing those skills outside of session, followed by the introduction of a new skill. Participants then engage in approximately 45 minutes of independent practice in their assigned experimental condition. Following independent practice, participants spend approximately 20 minutes debriefing with the therapist processing experiences and discussing how skills might be applied outside the experimental setting. Participants then complete post-session self-report measures. The conditions are designed to be closely matched on therapist time, session structure, and intervention content, while differing in the attentional focus of the mindfulness skills and the context in which skills are practiced. Participants in both conditions receive equivalent compensation, although total visit duration differs by condition: IFT sessions last approximately two and a half to three hours, whereas EFT sessions last approximately two to two and a half hours due to the additional procedural time required for changing, showering before and after floatation, and redressing.

### IFT and EFT Introduction Session

During the one-hour introduction session, the therapist introduces rationale for participant’s assigned condition, including nonjudgmental observation of experience. Additionally, both groups explore the connection between thoughts, emotions, and behaviors, including how these experiences relate to eating disorder and body image related concerns.

Psychoeducation is given on how each of these elements may increase engagement in disorder-specific behaviors. In IFT, focus on interoception and connection with internal body experience is also emphasized. Participants also complete two introductory mindfulness exercises tailored to condition. In IFT, participants practice a mindful breath exercise, directing attention to the internal bodily experience. In the EFT group, participants practice present-moment awareness that directs attention to an external focus. Figure 2 summarizes skills introduced and practiced in each condition.

**Figure 2.**
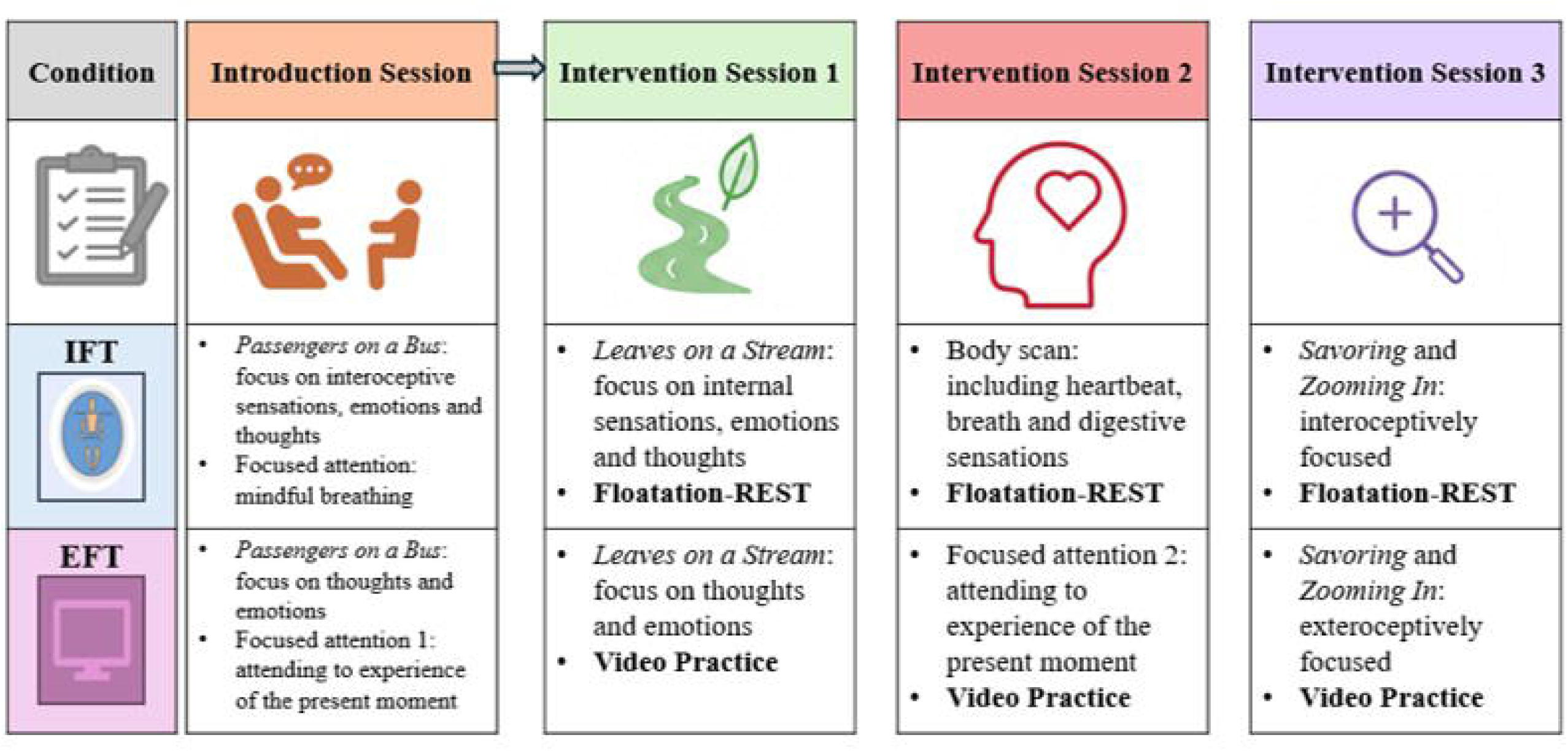
Skills Introduced During Experimental Sessions. *Note*. Figure depicts skills covered during the introduction and experimental sessions based on study condition. Bolded terms represent the skills practice respective to condition (i.e., Floatation-REST or Video Practice).

### ACT and MORE-Based Skills Used in IFT and EFT

Session protocols (developed by EMC) draw from principles presented in ACT (Hayes et al., 1999) and MORE (Garland, 2013). Protocols were adapted to distinguish an interoceptive versus exteroceptive attentional focus. See Supplementary A for information about discontinuation and treatment compliance.

ACT is a flexible approach with six core processes (i.e., cognitive defusion, acceptance, contact with the present moment, values, committed action, and self-as-context; Hayes et al., 1999). The present protocol primarily draws on contact with the present moment, acceptance, and cognitive defusion adapted according to the condition. In the IFT group, participants practice directing attention toward bodily sensations and observing these sensations along with associated thoughts and emotions. The aim is to increase acceptance of these sensations and reduce judgment. In contrast, the EFT condition applies parallel skills while directing attention toward the external environment and observing thoughts and emotions that arise without becoming disengaged from their present moment experience. For example, during the IFT body scan exercise, a participant may notice the experience of gastric sensations. Previously, this individual may associate these with negative thoughts (i.e., “I am hungry, if I eat, I am going to get fat”); however, the goal is that IFT helps the individual to experience and acknowledge the sensation without automatic evaluations (i.e., I notice I am having stomach rumbling). During the corresponding EFT exercise, the participant is directed to attend towards what it feels like to be present in one’s external environment. The participant notices sights and sounds and the experience of simply being while practicing nonjudgmental awareness of thoughts and emotions that arise.

Thought defusion and acceptance exercises are introduced in parallel across both conditions and include *Passengers on a Bus*, thought distancing, and *Leaves on a Stream*. In the IFT condition, participants are explicitly prompted to incorporate bodily sensations and judgments related to those sensations into these exercises. In the EFT condition, participants are not explicitly prompted to attend to bodily sensations. Participants may spontaneously report internal sensations; however, clinicians do not intentionally explore or elaborate on connections between those sensations, thoughts, and emotions.

*Savoring* and *Zooming In* are drawn from MORE (Garland, 2013). *Savoring* involves intentionally directing and sustaining attention towards pleasant features of an experience, with the aim of increasing awareness of positive and the potential reward of that experience. In both conditions, the exercise is intended to shift participants’ cognitions away from broad evaluative judgments and toward more specific features of their moment-to-moment experience. In IFT, participants apply this skill to their experience of their body, including neutral and pleasant sensations, with the goal of increasing awareness and acceptance. The ultimate goal is to enhance bodily trust through understanding and nonjudgment of their bodily experience. This emphasis is particularly relevant to AN, in which reduced trust and confidence in bodily sensations have been associated with greater eating-disorder psychopathology (Monteleone et al., 2021). The EFT condition applies the same principles to external sensory features, such as sights, textures, or colors. Participants bring an object with sentimental value to use during the exercise.

*Zooming In* is another strategy utilized to reduce broad evaluative interpretation of experiences to specific components and/or features. *Zooming In* involves shifting attention from broad evaluations of subjectively negative experiences toward better understanding and identification of the experience while also engaging in nonjudgement during the experience. For example, in the IFT group, participants apply this to internal body sensations with the goal of increasing differentiation between sensations and nonjudgmental awareness of the bodily experience rather than responding to sensations with global negative evaluations (e.g., “if I feel full, I am fat”). In EFT, this skill is used to break down a global negative evaluation about the environment and focus on how one unpleasant feature may make up a small part of the whole present experience.

### IFT Experimental Sessions

In addition to skills practice mentioned above, during the IFT experimental sessions, participants complete a 45-minute Floatation-REST session. During Floatation-REST exteroceptive input is reduced via multiple channels with the aim of enhancing attention to internal signals. Participants lay supine in one of two circular fiberglass pools that are light and soundproof, and are eight feet in diameter, containing ∼11 inches of water saturated with ∼1,800 pounds of Epsom salt (see Figure 3 and Supplementary A). This dense saltwater solution allows participants to effortlessly float while the water hovers just above the ears. This buoyance allows for reduced sensory information from gravity and proprioceptive input. The temperature of the water and air is calibrated to approximate the temperature of the skin (∼95.0 °F), helping to minimize the need for thermoregulation while reducing the boundary between air, body, and water. Unlike the picture in Figure 3, clothing is usually not worn while floating (to minimize somatosensory stimulation), though participants can choose to float with a bathing suit. In regard to interoceptive stimulation, previous work suggests Floatation-REST promotes increased attention to cardiorespiratory sensation (Feinstein et al., 2018a; Khalsa et al., 2020; Choquette et al., 2026). Further cardiorespiratory sensations were rated more positively during Floatation-REST condition compared to a comparator condition (Feinstein et al., 2018a; Khalsa et al., 2020; Choquette et al., 2026).

**Figure 3.** Floatation-REST Pools. *Note.* Floatation pools at the LIBR Float Clinic and Research Center. Left: Open pool; Right: Closed pool. Participants typically float in darkness, with the lights off. Figure is reproduced from https://doi.org/10.1371/journal.pone.0190292.g002 (Feinstein et al., 2018b).

### EFT Experimental Sessions

During the EFT experimental sessions, participants independently practice acceptance- and mindfulness-based skills using pre-recorded guided videos. During each approximately 45-minute practice period, participants view two to three videos that reinforce previously introduced skills and provide guided practice of the skill introduced during the current session. Participants are instructed to engage with and practice the targeted skill for the duration of each video.

Following each video, participants complete brief written reflections focused on their experience of the exercise, including their thoughts and emotions, ability to engage with the skill, and potential applications outside the session. Following completion of the video practice period, participants debrief their experience with the therapist.

### Training and Fidelity

Introduction and experimental sessions are currently delivered by the PI (EMC, a licensed psychologist). As enrollment progresses, sessions may also be delivered by a licensed doctoral or master’s level clinician or a therapist in training (i.e., clinical psychology post-doctoral fellow, pre-doctoral intern, or graduate student). Before delivering the study protocol, each therapist will complete training in the intervention protocol with the PI and review relevant manuals and supporting literature. The PI will also provide supervision to junior therapists throughout intervention delivery. Sessions are recorded for the purpose of assessing fidelity to treatment protocol. Fidelity ratings will be provided by consultants for at least 20% of intervention sessions to ensure the intervention quality and adherence to condition-specific protocol.

## Data Collection

To protect confidentiality, unique identification numbers are assigned to each participant. Hardcopy data is filed in locked office cabinets at LIBR. Electronic data (as well as video/audio recordings) are kept in a firewalled/password protected database on a secure server managed by LIBR and stored using identification numbers. Research Electronic Data Capture (REDCap; Harris et al., 2009) is used to collect self-report assessments. Behavioral tasks are hosted on StimTool3 (https://github.com/laureate-institute-for-brain-research/StimTool3), while psychophysiological data (i.e., heartbeat, respiration, and blood pressure) is recorded via BioPac MP150 system and AcqKnowledge software (BIOPAC Systems, Inc., Goleta, CA, USA).

Neuroimaging data is captured on a 3T Siemens MAGNETOM Prisma-NX magnetic resonance imaging (MRI; Siemens Healthineers, Malvern, PA, USA), and the platform Chorus (https://app.joinchorus.com/login) hosts the Somatomap tasks (Arevian et al., 2020). All web-based information transmission is encrypted and REDCap (Harris et al., 2009) servers are housed in a local data center at LIBR. Physiological, self-report, neuroimaging, and behavioral measures are taken throughout the study (see Table 1 for a complete list of measures and Supplementary A for further details).

### Measures

**Primary Outcome Measure.** The primary outcome is perceptual BID measured via the PFRS (Swami et al., 2008, 2012). On this measure, participants select the depiction of their current and ideal body from 10 images ranging from extremely emaciated (BMI = 12.51) to a larger body size (BMI = 41.23; Swami et al., 2008). BID is calculated as the absolute difference between current and ideal scores, with higher scores indicating greater perceptual BID. The primary outcomes are measured as acute change in perceptual BID from pre- to post-experimental session, as well as from baseline to post-intervention. Longer-term changes in PFRS scores assessed six-week and six-month follow-ups are secondary outcomes.

**Secondary Measures.** Other measures of perceptual BID include the Somatomap3D (during behavioral and functional magnetic resonance imaging (fMRI) sessions with concurrent scanning; Ralph-Nearman et al., 2019, 2021; Arevian et al., 2020) the String Task (Schneider et al., 2009), and the Aperture Task (Keizer et al., 2013; Irvine et al., 2019). To measure cognitive/affective BID, participants complete the Body Image State Scale (BISS; Cash et al., 2002) and the Multidimensional Body-Self Relations Questionnaire (MBSRQ; Cash, 2000). Self-report measures of interoception include the Multidimensional Assessment of Interoceptive Awareness (MAIA-V2; Mehling et al., 2012), the Interoceptive Attention Scale (IATS; Gabriele et al., 2022), the Somatomap2D Interoception (Ralph-Nearman et al., 2019, 2021; Arevian et al., 2020), and pre- and post-experimental session visual analogue scale (VAS) ratings of interoceptive sensations and bodily states (used in previous LIBR studies; Feinstein et al., 2018a, 2018b; Khalsa et al., 2020; Choquette et al., 2023). Behavioral tasks assessing interoception are the Heartbeat Tapping Task (HBT; Ludwick-Rosenthal and Neufeld, 1985; Canales-Johnson et al., 2015; Smith et al., 2021), the Inspiratory Breath Hold Task (Lapidus et al., 2020), and the Visceral Interoceptive Attention Task (VIA, completed during concurrent fMRI; Simmons et al., 2013).

Secondary outcomes for the Somatomap2D Interoception and VAS ratings are measured as acute changes from pre- to post-experimental session as well as from baseline to post-intervention. The BISS will be evaluated as acute within sessions changes. Changes from baseline to post-intervention will be evaluated in the following measures: Somatomap3D (behavioral and functional), String Task, Aperture Task, MBSRQ, MAIA-V2, IATS, HBT, Inspiratory Breath Hold Task, and VIA. Longer-term changes will be assessed at six-week and six-month follow-ups for the EDEQ, Somatomap3D, BISS, MBSRQ, MAIA-V2, and IATS.

## Data Analysis

Data will be investigated for missingness and attrition, and descriptive statistics will be calculated for all variables. All hypotheses will be tested at *α*=0.05 level, with appropriate adjustments for multiple testing. Significance levels and estimates of effect sizes will be provided for each analysis. For all models, relevant covariates will be included (e.g., age, body mass index, and eating disorder symptom severity measured at baseline). See Supplementary A for further details on analyses relating to secondary aims (testing the relationship with BID and interoception), and exploratory aims (to investigate longitudinal associations and mechanistic mediators of BID change in AN).

### Primary Aims, Hypotheses and Analyses

The primary aim is to examine how an interoceptive manipulation acutely changes BID in AN. We hypothesize that 1a) the IFT group will show greater perceptual BID (Swami et al., 2008; Schneider et al., 2009; Keizer et al., 2013; Irvine et al., 2019; Ralph-Nearman et al., 2019, 2021) reductions acutely and over the intervention period, compared to the EFT group. Alternatively, we hypothesize that 1b) cognitive/affective BID (Cash, 2000; Cash et al., 2002) reductions in the EFT group may be greater than the IFT group acutely and over the intervention period. Acute effects will be determined using linear mixed model (LMM) to explore condition (IFT versus EFT) by time (pre- to post-experimental session) interaction effects on perceptual BID (indexed by PFRS; Swami et al., 2008). Intervention effects will also be determined using LMM to explore a condition by time (baseline to post-intervention session) interaction effects on the PFRS.

Secondary analysis will include LMM to estimate effects of condition on self-report (Swami et al., 2008) and behavioral measures (Schneider et al., 2009; Keizer et al., 2013; Ralph-Nearman et al., 2019, 2021) from pre-intervention through completion of the post-intervention assessments. Condition, time, and their interaction will be modeled as fixed effects. LMM will be used to explore the condition by session interaction on cognitive/affective BID (indexed by the BISS; Cash et al., 2002) both acutely and across the intervention period (indexed by the MBRSQ; Cash, 2000).

Aim two is to determine cross-sectional multilevel associations between interoception and BID in AN. We hypothesize that 2a) perceptual and 2b) cognitive/affective BID will be associated with self-report (MAIA-V2; Mehling et al., 2012, IATS; Gabriele et al., 2022), interoceptive visual analogue scales (Feinstein et al., 2018a, 2018b; Khalsa et al., 2020; Choquette et al., 2023), and behavioral measures (HBT; Ludwick-Rosenthal and Neufeld, 1985; Canales-Johnson et al., 2015; Smith et al., 2021, Inspiratory Breath Hold task; Lapidus et al., 2020) of interoception. We also expect that 2c) perceptual BID will be more strongly related to interoception measures than cognitive/affective measures.

Exploratory aims include the longitudinal and mechanistic mediators of BID. For instance, 3a) group differences in perceptual BID (Swami et al., 2008; Ralph-Nearman et al., 2021) at six-week and six-month follow-up will be explored using LMM. Multilevel mediation models will additionally be fit to explore 3b) perceptual BID changes via multimodal interoception and state/trait illness measures of AN. Finally, we anticipate 3c) that BID will also be associated with neural activation in regions of interest (ROI) during the VIA (Simmons et al., 2013) task. Given its role in interoceptive processing (Wagner et al., 2008; Kaye et al., 2009; Kerr et al., 2016; Choquette and Khalsa, 2025b), we expect to see greater insular activation in the IFT condition during the interoceptive conditions compared to EFT condition.

### Sample Size

The sample size was determined via a power analysis simulation using R for the primary aim. A small effect size (*d* = 0.21) on PFRS (Swami et al., 2008). BID is expected based on a previous Floatation-REST intervention studies (Choquette et al., 2023). There is currently no data to estimate the exact effect of acceptance- and mindfulness-based interventions on perceptual BID, as prior clinical research in eating disorders has not included these outcome measures. Given that these techniques focus on cognitive aspects of body image rather than perceptual and enhancing interoceptive mindfulness and acceptance, we are conservatively estimating a smaller effect (*d* = 0.10) on perceptual BID. After accounting for a 10% dropout between baseline and post intervention, the current study would have 92 participants (46 per arm) and given simulations 80% power to detect a small effect within treatment arms and a small interaction effect (*d* > 0.20). A synergistic effect of *d* = 0.31 is expected such that float combined with IFT will show greater improvements on BID for both within treatment arm and interaction effects. Thus, we will be well-powered for within subject effects and adequately powered for estimated interaction effects.

### Gender/Minority/Pediatric Inclusion

This study enrolls only female participants which reflects primarily the predominance of AN in females (10:1 female to male ratio; American Psychiatric Association, 2022) Further considerations include that the target sample size is underpowered to detect sex differences and recruitment site only enrolls biologically female patients. Given the current protocol is believed to pose minimal risk to children, and previous Floatation-REST studies have included children as young as 13 with no adverse effects (Khalsa et al., 2020; Choquette et al., 2023). This study involves volunteer subjects 13-17 years old. This is an important developmental period given the age of AN onset is most frequently in adolescence (Klein and Walsh, 2003). Finally, every effort will be made to recruit minority participants; given the demographic makeup of the recruitment site, it is expected participants will primarily be Non-Hispanic White.

### Safety

The current protocol is thought to have minimal added risk. As the study population is at increased risk for negative medical and mental health events, safety and monitoring will be carried out by PI (EMC) and independent safety monitor (ISM), with consultation from the grant mentorship team comprised of experts in the field. Additionally, adverse events (AEs) are promptly communicated to the LIBR Human Protection Administrator (HPA) to determine the necessity of immediate IRB report. All AEs are included in the IRB annual report. Medical risk/relapses are overseen while participants are in treatment by their LEDP providers. Suicide risk is monitored over the course of the study by both LEDP providers during treatment and by research staff. If needed, safety assessments and/or safety plans will be conducted with a staff member, and referrals may be provided to the LEDP psychiatrist, local medical provider, or the 24-hour assessment at department on the same campus as LIBR at the Laureate Psychiatric Clinic and Hospital (see Supplementary A for further information). If a participant is no longer in the Tulsa Metropolitan area specific resources to their location will be provided. Unanticipated, serious adverse events will be reported to the LIBR HPA, IRB, and ISM within 24 hours and the National Institutes of Health (NIH) Program Officer within 48 hours of the time project staff become aware of the incident.

### Dissemination

Data from this project will be shared with the scientific community through presentation of results at scientific meetings focused on eating disorders, interoception, contemplative practices, and psychophysiology, and publication in scientific journals. All final peer reviewed manuscripts will be submitted to the digital archive PubMed Central, and we will maintain a local website (at www.laureateinstitute.org) where project information and lay summaries of scientific presentations and publications will be posted. Additionally, results will be submitted to ClinicalTrials.gov.

## Discussion

Given the recalcitrant nature of BID in AN (Chapa et al., 2020), there is great imperative toward understanding how targetable processes, such as interoception, relates to BID in AN. The current protocol provides a framework of integrating Floatation-REST and skills from third-wave therapies, while leveraging longitudinal data collection to probe potential mechanisms behind change in BID.

The present study is unique in using multimodal assessments to examine the role of interoception on BID in AN. BID and interoception have traditionally been assessed via self-report questionnaires. While self-report provides valuable insights into subjective elements of these constructs, emerging behavioral tasks allow for objective measures, enabling a more holistic perspective. For BID, behavioral tasks permit actual body image distortion to be quantified when taking the difference between estimated (i.e., string; Schneider et al., 2009, aperture task; Keizer et al., 2013; Irvine et al., 2019) and true body size as taken by anthropometric body measures. Additionally, cardiorespiratory interoception can be probed with the Heartbeat Tapping (Ludwick-Rosenthal and Neufeld, 1985; Canales-Johnson et al., 2015; Smith et al., 2021) and Inspiratory Breath Hold Tasks (Lapidus et al., 2020), which introduce holding the breath as an interoceptive disruption for one’s system and task performance to adapt to.

Beyond the addition of behavioral assessments, this protocol harnesses neural measures designed to precisely assess networks of interoceptive processing that spans multiple biological systems (i.e., cardiovascular, gastrointestinal, and respiratory), as well as the perceptual and cognitive/affective facets of BID. The inclusion of functional neuroimaging presents an important step toward elucidating neural mechanisms of BID in AN and exploring the relationship between BID and interoception. It is posited that abnormalities in body image processing may arise from difficulties in calibrating incoming ‘bottom up’ perceptual information with an excessive attention to ‘top down’ cognitive schemas and valence attributions (Diaz-Fong and Feusner, 2024). In the current study, the VIA is used to probe neural responses associated with top-down processing of interoception. Additionally, the Somatomap3D is used to explore processing of perceptual body image. Examining changes in responses following IFT and EFT may provide insight into whether intervention-related reductions in BID or improvements in interoception are accompanied by changes in neural processing of body-related information.

Importantly, with the high relapse rates imbued in AN following standard treatment (Steinhausen, 2002; Khalsa et al., 2017), results from the study’s mindfulness-based interventions may allow for potentially scalable and non-invasive ancillary treatment avenues. This study is innovative in the inclusion of elements to specifically address both the cognitive/affective and perceptual components of BID. In turn, the current interventions may allow for precision medicine approaches depending on the facet of BID demanding the most immediate therapeutic focus. Interoception may represent a similarly individually adaptable target via mindfulness-based practices such as Floatation-REST, the acceptability of which has been initially established in AN (Khalsa et al., 2020). Taken together, results from this study may pave the way for the expansion of these mindfulness-based interventions for interoception and BID that are safe and feasible for treatment enhancement.

The current protocol is not without limitations. Generalizability is limited by the restriction of recruitment to a single treatment center and to women and girls. Future work may expand recruitment to multiple sites to improve both generalizability of results and statistical power.

Additionally, this study does not include a comparator REST condition. A third arm was considered wherein participants engage in an active interoceptive comparator such as REST via antigravity chair or Floatation-REST alone. However, as the purpose of the current project is to determine the effects of interoceptive manipulation on BID, EFT provides an exteroceptively focused comparison, aspects of which have had clinical impacts on cognitive/affective aspects of BID. Future studies may include alternative forms of REST to determine if the effect is unique to Floatation-REST. Further, while the current protocol is limited to AN, subsequent work may explore the effects of interoceptive manipulation on BID in relevant psychiatric conditions including bulimia nervosa and body dysmorphic disorder.

Overall, this protocol takes a systematic approach to explore the relationship with BID and interoception, and proposes mindfulness-based mechanistic treatments for BID in AN. As a proof-of-concept trial, additional steps could include full scale mindfulness-based intervention development for AN as well as other BID-related psychiatric conditions. Taken together, knowledge gained from this study may facilitate novel approaches to BID via modulation of interoception as a target.

## Trial Status

The study received approval from the Western Institutional Review Board (WCG IRB; 20234215) on 9/26/2023, and the current protocol is Version 5.0 (see Supplementary A Table 1 for protocol versions and amendments). Recruitment for the trial started approximately on 4/1/2024, with an estimated completion date of 11/1/2028. 80 participants have consented at the time of manuscript submission The study was registered on ClinicalTrials.gov (NCT06332963) on 3/27/2024.

## Supplementary Material

Supplementary A. Further information of methods of intervention, assessments, and analyses.

Supplementary B: completed SPIRIT checklist.

Supplementary C: initial study approval letter from the Western Institutional Review Board (WCG IRB).

Supplementary D: K23 Notice of Award: letter to the principal investigator from the National Institute of Mental Health with notice of the award for the current study.

Supplementary E: WCG IRB approved consent form.

## Abbreviations

ACT: Acceptance and Commitment Therapy
AEs: Adverse Events
AN: Anorexia Nervosa
ARFID: Avoidant/Restrictive Food Intake Disorder Screen
ASI: Anxiety Sensitivity Inventory
BBDS: Body Boundary Dissolution Scale
BIAAQ: Body Image Acceptance and Action Questionnaire
BID: Body Image Dissatisfaction
BISS: Body Image State Scale
BMI: Body Mass Index
CEQ: Credibility/Expectancies Questionnaire
COPES: Community Outreach Psychiatric Emergency Services
C-SSRS: Columbia-Suicide Severity Rating Scale
DSM: Diagnostic and Statistical Manual of Mental Disorders
ECG: Electrocardiogram
EDE-Q: Eating Disorder Examination Questionnaire
EDI: Eating Disorder Inventory
EFT: Exteroceptively Focused Treatment
FCRC: Float Clinic and Research Center
fMRI: functional Magnetic Resonance Imaging
GSQ: Glasgow Sensory Questionnaire
HBT: Heartbeat Tapping Task
HPA: Human Protection Administrator
IATS: Interoceptive Attention Scale
IFT: Interoceptively Focused Treatment
KSS: Karolinska Sleepiness Scale
LED: Light-Emitting Diode
LEDP: Laureate Eating Disorders Program
LIBR: Laureate Institute for Brain Research
LLM: Linear Mixed Model
IRB: Institutional Review Board
ISM: Independent Safety Monitor
MAIA: Multidimensional Assessment of Interoceptive Awareness
MBSRQ: Multidimensional Body-Self Relations Questionnaire
MORE: Mindfulness-Oriented Recovery Enhancement
NIH: National Institutes of Health
PANAS: Positive and Negative Affect Schedule
PFRS: Photographic Figure Rating Scale
PI: Principal Investigator
PHQ: Patient Health Questionnaire
PLUS: Participant Last Use Summary
REDCap: Research Electronic Data Capture
REST: Reduced Environmental Stimulation Therapy
ROI: Regions of Interest
SAE: Serious Adverse Event
STAI: State Trait Anxiety Inventory
UV: Ultraviolet
VIA: Visceral Interoceptive Attention Task
VSI: Visceral Sensitivity Index
WAI-SR: Working Alliance Inventory – Self Report
WCG IRB: Western Institutional Review Board
5D-ASC: 5 Dimension Altered States of Consciousness Scale

## Data Availability

Deidentified data from this ongoing trial have been submitted to the National Institute of Mental Health Data Archive (NDA; project ID: C5116; https://doi.org/10.15154/7c0r-rt56). Data will be released upon study completion in accordance with the approved data sharing plan. Access to released participant-level data will be subject to NDA approval and applicable data use requirements. No study results are reported in this protocol manuscript.

## Acknowledgements

The authors would like to acknowledge Drs. Sahib S. Khalsa, Robin L., Aupperle, and Nancy L. Zucker for their mentorship throughout the K23 application and study period and contributions to the development of the therapy protocols. They would also like to acknowledge Abhinita Premkumar and Rachel Thompson who were instrumental in data collection and study coordination, Dr. Eric Garland for sharing expertise and protocols from Mindfulness-Oriented Recovery Enhancement for this study. Finally, the authors would like to thank Armen Arevian and Jamie Feusner for their assistance with task initialization and Matt Mann, MWP who graciously allowed for the use of his videos for the Leaves on a Stream exercise (https://manns-woodland-perspective.com/).

## Generative AI statement

During the preparation of this manuscript, authors used generative AI tool (ChatGPT; OpenAI) to assist with language editing, clarity, organization, and refinement of wording. The tool was not used to generate content or make scientific conclusions. All AI-assisted content was critically reviewed and revised by authors for accuracy. The authors take responsibility for accuracy, integrity, and final content of the manuscript.

## CRediT Authorship Contribution Statement

**Elizabeth Bernardini:** Writing-original draft, Writing-review and editing, Visualization, Investigation. **Emily Choquette:** Writing-original draft, Writing-review and editing, Conceptualization, Methodology, Investigation, Visualization, Supervision, Project administration, Funding Acquisition.

## References

Åkerstedt, T., and Gillberg, M. (1990). Subjective and objective sleepiness in the active individual. Int. J. Neurosci. 52, 29–37. doi: 10.3109/00207459008994241

Alleva, J. M., Sheeran, P., Webb, T. L., Martijn, C., and Miles, E. (2015). A meta-analytic review of stand-alone interventions to improve body image. PLoS One 10, e0139177. doi: 10.1371/journal.pone.0139177

American Psychiatric Association (2022). Diagnostic and Statistical Manual of Mental Disorders. DSM-5-TR. Washington, DC: American Psychiatric Association Publishing. doi: 10.1176/appi.books.9780890425787

Arevian, A. C., O’Hora, J., Rosser, J., Mango, J. D., Miklowitz, D. J., and Wells, K. B. (2020). Patient and provider cocreation of mobile texting apps to support behavioral health: usability study. JMIR mHealth uHealth 8, e12655. doi: 10.2196/12655

Canales-Johnson, A., Silva, C., Huepe, D., Rivera-Rei, Á., Noreika, V., Garcia, M. del C., et al. (2015). Auditory feedback differentially modulates behavioral and neural markers of objective and subjective performance when tapping to your heartbeat. Cereb. Cortex 25, 4490–4503. doi: 10.1093/cercor/bhv076

Carter, J. C., Blackmore, E., Sutandar-Pinnock, K., and Woodside, D. B. (2004). Relapse in anorexia nervosa: a survival analysis. Psychol. Med. 34, 671–679. doi: 10.1017/S0033291703001168

Carter, J. C., Mercer-Lynn, K. B., Norwood, S. J., Bewell-Weiss, C. V., Crosby, R. D., Woodside, D. B., et al. (2012). A prospective study of predictors of relapse in anorexia nervosa: implications for relapse prevention. Psychiatry Res. 200, 518–523. doi: 10.1016/j.psychres.2012.04.037

Cash, T. F. (2000). The Multidimensional Body-Self Relations Questionnaire Users’ Manual. Norfolk, VA: Old Dominion University.

Cash, T. F. (2004). Body image: past, present, and future. Body Image 1, 1–5. doi: 10.1016/S1740-1445(03)00011-1

Cash, T. F., and Deagle, E. A. (1997). The nature and extent of body-image disturbances in anorexia nervosa and bulimia nervosa: a meta-analysis. Int. J. Eat. Disord. 22, 107–125. doi: 10.1002/(SICI)1098-108X(199709)22:2<107::AID-EAT1>3.0.CO;2-J

Cash, T. F., Fleming, E. C., Alindogan, J., Steadman, L., and Whitehead, A. (2002). Beyond body image as a trait: the development and validation of the Body Image States Scale. Eat. Disord. 10, 103–113. doi: 10.1080/10640260290081678

Cash, T. F., and Green, G. K. (1986). Body weight and body image among college women: perception, cognition, and affect. J. Pers. Assess. 50, 290–301. doi: 10.1207/s15327752jpa5002_15

Cash, T. F., and Smolak, L. (Eds.) (2012). Body Image: A Handbook of Science, Practice, and Prevention. 2nd Edn. New York, NY: Guilford Press.

Chan, A.-W., Boutron, I., Hopewell, S., Moher, D., Schulz, K. F., Collins, G. S., et al. (2025). SPIRIT 2025 statement: updated guideline for protocols of randomised trials. BMJ 389, e081477. doi: 10.1136/bmj-2024-081477

Chapa, D. A. N., Hagan, K. E., Forbush, K. T., Clark, K. E., Tregarten, J. P., and Argue, S. (2020). Longitudinal trajectories of behavior change in a national sample of patients seeking eating-disorder treatment. Int. J. Eat. Disord. 53, 917–925. doi: 10.1002/eat.23272

Choquette, E. M., Flux, M. C., Moseman, S. E., Chappelle, S., Naegele, J., Upshaw, V., et al. (2023). The impact of floatation therapy on body image and anxiety in anorexia nervosa: a randomised clinical efficacy trial. EClinicalMedicine 64, 102173. doi: 10.1016/j.eclinm.2023.102173

Choquette, E. M., Garland, M., Morrissey, G., Wilzok, N., Wilson, R., Premkumar, A., et al. (2026). Reduced Environmental Stimulation Therapy (REST) in methamphetamine use disorder: a pilot study. Addict. Behav. Rep. 23, 100651. doi: 10.1016/j.abrep.2025.100651

Choquette, E. M., and Khalsa, S. S. (2025a). “Interoception in Eating Disorders,” in The Handbook of the Neurobiology of Eating Disorders, eds. C. E. Wierenga and J. E. Steinglass (New York, NY: Oxford University Press), 162–188. doi: 10.1093/9780197675212.003.0009

Choquette, E. M., and Khalsa, S. S. (2025b). Sensing the self: the role of the insula and interoception in body image. Curr. Top. Behav. Neurosci. 70, 107–125. doi: 10.1007/7854_2025_581

Dambrun, M. (2016). When the dissolution of perceived body boundaries elicits happiness: the effect of selflessness induced by a body scan meditation. Conscious. Cogn. 46, 89–98. doi: 10.1016/j.concog.2016.09.013

Delinsky, S. S., and Wilson, G. T. (2006). Mirror exposure for the treatment of body image disturbance. Int. J. Eat. Disord. 39, 108–116. doi: 10.1002/eat.20207

Deloitte Access Economics (2020). Social and economic cost of eating disorders in the United States of America. https://www.hsph.harvard.edu/striped/report-economic-costs-of-eating-disorders/ [Accessed October 20, 2026].

Devilly, G. J., and Borkovec, T. D. (2000). Psychometric properties of the credibility/expectancy questionnaire. J. Behav. Ther. Exp. Psychiatry 31, 73–86. doi: 10.1016/s0005-7916(00)00012-4

Diaz-Fong, J. P., and Feusner, J. D. (2024). “Visual Perceptual Processing Abnormalities in Body Dysmorphic Disorder,” in Perceptual Dysregulation in Psychiatric Nosology, eds. S. Khalsa and A. Powers (Cham: Springer Nature Switzerland), 105–127. doi: 10.1007/7854_2024_472

Fairburn, C. G. (2008). Cognitive Behavior Therapy and Eating Disorders. New York: Guilford Press.

Feinstein, J. S., Khalsa, S. S., Yeh, H., Al Zoubi, O., Arevian, A. C., Wohlrab, C., et al. (2018a). The elicitation of relaxation and interoceptive awareness using floatation therapy in individuals with high anxiety sensitivity. Biol. Psychiatry Cogn. Neurosci. Neuroimaging 3, 555–562. doi: 10.1016/j.bpsc.2018.02.005

Feinstein, J. S., Khalsa, S. S., Yeh, H., Wohlrab, C., Simmons, W. K., Stein, M. B., et al. (2018b). Examining the short-term anxiolytic and antidepressant effect of Floatation-REST. PLoS One 13, e0190292. doi: 10.1371/journal.pone.0190292

Fischer, D., Messner, M., and Pollatos, O. (2017). Improvement of interoceptive processes after an 8-week body scan intervention. Front. Hum. Neurosci. 11, 452. doi: 10.3389/fnhum.2017.00452

Fredrickson, B. L., and Roberts, T. (1997). Objectification theory: toward understanding women’s lived experiences and mental health risks. Psychol. Women Q. 21, 173–206. doi: 10.1111/j.1471-6402.1997.tb00108.x

Gabriele, E., Spooner, R., Brewer, R., and Murphy, J. (2022). Dissociations between self-reported interoceptive accuracy and attention: evidence from the Interoceptive Attention Scale. Biol. Psychol. 168, 108243. doi: 10.1016/j.biopsycho.2021.108243

Gadsby, S. (2017). Distorted body representations in anorexia nervosa. Conscious. Cogn. 51, 17–33. doi: 10.1016/j.concog.2017.02.015

Galmiche, M., Déchelotte, P., Lambert, G., and Tavolacci, M. P. (2019). Prevalence of eating disorders over the 2000-2018 period: a systematic literature review. Am. J. Clin. Nutr. 109, 1402–1413. doi: 10.1093/ajcn/nqy342

Gardner, R. M., and Brown, D. L. (2014). Body size estimation in anorexia nervosa: a brief review of findings from 2003 through 2013. Psychiatry Res. 219, 407–410. doi: 10.1016/j.psychres.2014.06.029

Garfinkel, P. E., Moldofsky, H., Garner, D. M., Stancer, H. C., and Coscina, D. V. (1978). Body awareness in anorexia nervosa: disturbances in “body image” and “satiety.” Psychosom. Med. 40, 487–498. doi: 10.1097/00006842-197810000-00004

Garland, E. L. (2013). Mindfulness-Oriented Recovery Enhancement for Addiction, Stress, and Pain. 1st Edn. Washington, DC: National Association of Social Workers (NASW) Press.

Garland, E. L. (2024). Mindfulness-Oriented Recovery Enhancement: implementing an evidence-based intervention for chronic pain, opioid use, and opioid addiction in clinical settings. Br. J. Clin. Pharmacol. 90, 3028–3035. doi: 10.1111/bcp.16147

Garner, D. M. (1991). Eating Disorder Inventory-2: Professional Manual. Odessa, FL: Psychological Assessment Resources.

Gopan, H., Rajkumar, E., Gopi, A., and Romate, J. (2024). Mindfulness-based interventions for body image dissatisfaction among clinical population: a systematic review and meta-analysis. Br. J. Health Psychol. 29, 488–509. doi: 10.1111/bjhp.12710

Griffen, T. C., Naumann, E., and Hildebrandt, T. (2018). Mirror exposure therapy for body image disturbances and eating disorders: a review. Clin. Psychol. Rev. 65, 163–174. doi: 10.1016/j.cpr.2018.08.006

Harris, E. C., and Barraclough, B. (1998). Excess mortality of mental disorder. Br. J. Psychiatry 173, 11–53. doi: 10.1192/bjp.173.1.11

Harris, P. A., Taylor, R., Thielke, R., Payne, J., Gonzalez, N., and Conde, J. G. (2009). Research electronic data capture (REDCap)—a metadata-driven methodology and workflow process for providing translational research informatics support. J. Biomed. Inform. 42, 377–381. doi: 10.1016/j.jbi.2008.08.010

Hayes, S. C., Strosahl, K., and Wilson, K. G. (1999). Acceptance and Commitment Therapy: An Experiential Approach to Behavior Change. New York, NY: Guilford Press.

Hoang, U., Goldacre, M., and James, A. (2014). Mortality following hospital discharge with a diagnosis of eating disorder: national record linkage study, England, 2001-2009. Int. J. Eat. Disord. 47, 507–515. doi: 10.1002/eat.22249

Horvath, A. O., and Greenberg, L. S. (1989). Development and validation of the Working Alliance Inventory. J. Couns. Psychol. 36, 223–233. doi: 10.1037/0022-0167.36.2.223

Irvine, K. R., McCarty, K., McKenzie, K. J., Pollet, T. V., Cornelissen, K. K., Tovée, M. J., et al. (2019). Distorted body image influences body schema in individuals with negative bodily attitudes. Neuropsychologia 122, 38–50. doi: 10.1016/j.neuropsychologia.2018.11.015

Kaye, W. H., Fudge, J. L., and Paulus, M. (2009). New insights into symptoms and neurocircuit function of anorexia nervosa. Nat. Rev. Neurosci. 10, 573–584. doi: 10.1038/nrn2682

Keel, P. K., Dorer, D. J., Franko, D. L., Jackson, S. C., and Herzog, D. B. (2005). Postremission predictors of relapse in women with eating disorders. Am. J. Psychiatry 162, 2263–2268. doi: 10.1176/appi.ajp.162.12.2263

Keizer, A., Smeets, M. A. M., Dijkerman, H. C., Uzunbajakau, S. A., van Elburg, A., and Postma, A. (2013). Too fat to fit through the door: first evidence for disturbed body-scaled action in anorexia nervosa during locomotion. PLoS One 8, e64602. doi: 10.1371/journal.pone.0064602

Kerr, K. L., Moseman, S. E., Avery, J. A., Bodurka, J., Zucker, N. L., and Simmons, W. K. (2016). Altered insula activity during visceral interoception in weight-restored patients with anorexia nervosa. Neuropsychopharmacology 41, 521–528. doi: 10.1038/npp.2015.174

Khalsa, S. S., Moseman, S. E., Yeh, H., Upshaw, V., Persac, B., Breese, E., et al. (2020). Reduced Environmental Stimulation in anorexia nervosa: an early-phase clinical trial. Front. Psychol. 11, 567499. doi: 10.3389/fpsyg.2020.567499

Khalsa, S. S., Portnoff, L. C., McCurdy-McKinnon, D., and Feusner, J. D. (2017). What happens after treatment? A systematic review of relapse, remission, and recovery in anorexia nervosa. J. Eat. Disord. 5, 20. doi: 10.1186/s40337-017-0145-3

Klein, D. A., and Walsh, B. T. (2003). Eating disorders. Int. Rev. Psychiatry 15, 205–216. doi: 10.1080/0954026031000136839

Kroenke, K., Spitzer, R. L., and Williams, J. B. (2001). The PHQ-9: validity of a brief depression severity measure. J. Gen. Intern. Med. 16, 606–613. doi: 10.1046/j.1525-1497.2001.016009606.x

Labus, J. S., Bolus, R., Chang, L., Wiklund, I., Naesdal, J., Mayer, E. A., et al. (2004). The Visceral Sensitivity Index: development and validation of a gastrointestinal symptom-specific anxiety scale. Aliment. Pharmacol. Ther. 20, 89–97. doi: 10.1111/j.1365-2036.2004.02007.x

Lapidus, R. C., Puhl, M., Kuplicki, R., Stewart, J. L., Paulus, M. P., Rhudy, J. L., et al. (2020). Heightened affective response to perturbation of respiratory but not pain signals in eating, mood, and anxiety disorders. PLoS One 15, e0235346. doi: 10.1371/journal.pone.0235346

Lima-Araujo, G. L. de, de Sousa Júnior, G. M., Mendes, T., Demarzo, M., Farb, N., Barros de Araujo, D., et al. (2022). The impact of a brief mindfulness training on interoception: a randomized controlled trial. PLoS One 17, e0273864. doi: 10.1371/journal.pone.0273864

Linardon, J., Gleeson, J., Yap, K., Murphy, K., and Brennan, L. (2019). Meta-analysis of the effects of third-wave behavioural interventions on disordered eating and body image concerns: implications for eating disorder prevention. Cogn. Behav. Ther. 48, 15–38. doi: 10.1080/16506073.2018.1517389

Lisdahl, K. M., Sher, K. J., Conway, K. P., Gonzalez, R., Feldstein Ewing, S. W., Nixon, S. J., et al. (2018). Adolescent brain cognitive development (ABCD) study: overview of substance use assessment methods. Dev. Cogn. Neurosci. 32, 80–96. doi: 10.1016/j.dcn.2018.02.007

Ludwick-Rosenthal, R., and Neufeld, R. W. (1985). Heart beat interoception: a study of individual differences. Int. J. Psychophysiol. 3, 57–65. doi: 10.1016/0167-8760(85)90020-0

Matthews, S. C., Simmons, A. N., Strigo, I. A., Arce, E., Stein, M. B., and Paulus, M. P. (2010). Escitalopram attenuates posterior cingulate activity during self-evaluation in healthy volunteers. Psychiatry Res. 182, 81–87. doi: 10.1016/j.pscychresns.2010.02.003

Mehling, W. E., Price, C., Daubenmier, J. J., Acree, M., Bartmess, E., and Stewart, A. (2012). The Multidimensional Assessment of Interoceptive Awareness (MAIA). PLoS One 7, e48230. doi: 10.1371/journal.pone.0048230

Mölbert, S. C., Klein, L., Thaler, A., Mohler, B. J., Brozzo, C., Martus, P., et al. (2017). Depictive and metric body size estimation in anorexia nervosa and bulimia nervosa: a systematic review and meta-analysis. Clin. Psychol. Rev. 57, 21–31. doi: 10.1016/j.cpr.2017.08.005

Monteleone, A. M., Cascino, G., Martini, M., Patriciello, G., Ruzzi, V., Delsedime, N., et al. (2021). Confidence in one-self and confidence in one’s own body: the revival of an old paradigm for anorexia nervosa. Clin. Psychol. Psychother. 28, 818–827. doi: 10.1002/cpp.2535

Porras Garcia, B., Ferrer Garcia, M., Olszewska, A., Yilmaz, L., González Ibañez, C., Gracia Blanes, M., et al. (2019). Is this my own body? Changing the perceptual and affective body image experience among college students using a new virtual reality embodiment-based technique. J. Clin. Med. 8, 925. doi: 10.3390/jcm8070925

Posner, K. (2016). Columbia-Suicide Severity Rating Scale. APA PsycTests. doi: 10.1037/t52667-000

Preston, C., and Ehrsson, H. H. (2014). Illusory changes in body size modulate body satisfaction in a way that is related to non-clinical eating disorder psychopathology. PLoS One 9, e85773. doi: 10.1371/journal.pone.0085773

Ralph-Nearman, C., Arevian, A. C., Moseman, S., Sinik, M., Chappelle, S., Feusner, J. D., et al. (2021). Visual mapping of body image disturbance in anorexia nervosa reveals objective markers of illness severity. Sci. Rep. 11, 12262. doi: 10.1038/s41598-021-90739-w

Ralph-Nearman, C., Arevian, A. C., Puhl, M., Kumar, R., Villaroman, D., Suthana, N., et al. (2019). A novel mobile tool (Somatomap) to assess body image perception pilot tested with fashion models and nonmodels: cross-sectional study. JMIR Ment. Health 6, e14115. doi: 10.2196/14115

Riva, G., and Gaudio, S. (2012). Allocentric lock in anorexia nervosa: new evidences from neuroimaging studies. Med. Hypotheses 79, 113–117. doi: 10.1016/j.mehy.2012.03.036

Riva, G., Malighetti, C., and Serino, S. (2021). Virtual reality in the treatment of eating disorders. Clin. Psychol. Psychother. 28, 477–488. doi: 10.1002/cpp.2622

Roberts, R. L., Ledermann, K., and Garland, E. L. (2022). Mindfulness-oriented recovery enhancement improves negative emotion regulation among opioid-treated chronic pain patients by increasing interoceptive awareness. J. Psychosom. Res. 152, 110677. doi: 10.1016/j.jpsychores.2021.110677

Robertson, A. E., and Simmons, D. R. (2013). The relationship between sensory sensitivity and autistic traits in the general population. J. Autism Dev. Disord. 43, 775–784. doi: 10.1007/s10803-012-1608-7

Rosen, J. C. (1996). Body image assessment and treatment in controlled studies of eating disorders. Int. J. Eat. Disord. 20, 331–343. doi: 10.1002/(SICI)1098-108X(199612)20:4<331::AID-EAT1>3.0.CO;2-O

Sandoz, E. K., Wilson, K. G., Merwin, R. M., and Kellum, K. (2013). Assessment of body image flexibility: the Body Image-Acceptance and Action Questionnaire. J. Contextual Behav. Sci. 2, 39–48. doi: 10.1016/j.jcbs.2013.03.002

Schneider, N., Frieler, K., Pfeiffer, E., Lehmkuhl, U., and Salbach-Andrae, H. (2009). Comparison of body size estimation in adolescents with different types of eating disorders. Eur. Eat. Disord. Rev. 17, 468–475. doi: 10.1002/erv.956

Simmons, W. K., Avery, J. A., Barcalow, J. C., Bodurka, J., Drevets, W. C., and Bellgowan, P. (2013). Keeping the body in mind: insula functional organization and functional connectivity integrate interoceptive, exteroceptive, and emotional awareness. Hum. Brain Mapp. 34, 2944–2958. doi: 10.1002/hbm.22113

Smith, R., Feinstein, J. S., Kuplicki, R., Forthman, K. L., Stewart, J. L., Paulus, M. P., et al. (2021). Perceptual insensitivity to the modulation of interoceptive signals in depression, anxiety, and substance use disorders. Sci. Rep. 11, 2108. doi: 10.1038/s41598-021-81307-3

Spielberger, C., Gorsuch, R., and Lushene, R. (1970). Manual for the State-Trait Anxiety Inventory. Palo Alto, CA: Consulting Psychologists Press.

Stanghellini, G., Ballerini, M., and Mancini, M. (2019). The optical-coenaesthetic disproportion hypothesis of feeding and eating disorders in the light of neuroscience. Front. Psychiatry 10, 630. doi: 10.3389/fpsyt.2019.00630

Steinhausen, H.-C. (2002). The outcome of anorexia nervosa in the 20th century. Am. J. Psychiatry 159, 1284–1293. doi: 10.1176/appi.ajp.159.8.1284

Studerus, E., Gamma, A., and Vollenweider, F. X. (2010). Psychometric evaluation of the Altered States of Consciousness Rating Scale (OAV). PLoS One 5, e12412. doi: 10.1371/journal.pone.0012412

Swami, V., Salem, N., Furnham, A., and Tovée, M. J. (2008). Initial examination of the validity and reliability of the female photographic figure rating scale for body image assessment. Pers. Individ. Dif. 44, 1752–1761. doi: 10.1016/j.paid.2008.02.002

Swami, V., Stieger, S., Harris, A. S., Nader, I. W., Pietschnig, J., Voracek, M., et al. (2012). Further investigation of the validity and reliability of the photographic figure rating scale for body image assessment. J. Pers. Assess. 94, 404–409. doi: 10.1080/00223891.2012.660293

Taylor, S., Zvolensky, M. J., Cox, B. J., Deacon, B., Heimberg, R. G., Ledley, D. R., et al. (2007). Robust dimensions of anxiety sensitivity: development and initial validation of the Anxiety Sensitivity Index-3. Psychol. Assess. 19, 176–188. doi: 10.1037/1040-<otherinfo> 3</otherinfo>590.19.2.176

Wagner, A., Aizenstein, H., Mazurkewicz, L., Fudge, J., Frank, G. K., Putnam, K., et al. (2008). Altered insula response to taste stimuli in individuals recovered from restricting-type anorexia nervosa. Neuropsychopharmacology 33, 513–523. doi: 10.1038/sj.npp.1301443

Watson, D., Clark, L. A., and Tellegen, A. (1988). Development and validation of brief measures of positive and negative affect: the PANAS scales. J. Pers. Soc. Psychol. 54, 1063–1070. doi: 10.1037//0022-3514.54.6.1063

Zickgraf, H. F., and Ellis, J. M. (2018). Initial validation of the Nine Item Avoidant/Restrictive Food Intake disorder screen (NIAS): a measure of three restrictive eating patterns. Appetite 123, 32–42. doi: 10.1016/j.appet.2017.11.111

Ziser, K., Mölbert, S. C., Stuber, F., Giel, K. E., Zipfel, S., and Junne, F. (2018). Effectiveness of body image directed interventions in patients with anorexia nervosa: a systematic review. Int. J. Eat. Disord. 51, 1121–1127. doi: 10.1002/eat.22946

