## Supplementary A for "Interoceptive mechanisms of body image disturbance in anorexia nervosa: Study protocol of a randomized parallel-group trial"

### Supplementary Methods

#### Recruitment and Retention

##### *Recruitment and Screening*

Initial recruitment and screening take place via referral from Laureate Eating Disorders Program (LEDP) physicians, therapists, nurses, or other clinical staff during weekly staffing meetings, verbally, or via email. First, providers confirm that to their knowledge, the participant meets the preliminary inclusion criteria, then the study staff contacts the participant and offers the opportunity to discuss study details during a consent visit.

After consent, participants provide a release of information for medical record access for one year, and complete initial screening (i.e., Columbia-Suicide Severity Rating Scale (C-SSRS; Posner, 2016), psychiatric and medical history via medical chart review, and self-reports). Urine drug and pregnancy screen and breathalyzer are conducted during one of the baseline visits prior to magnetic resonance imaging (MRI) scanning. Participants who do not meet eligibility criteria that may change over time, such as blood pressure requirements, may be rescreened in 7 days. If recent skin-lacerating self-harm is reported by the participant during the C-SSRS at screening, the participant may be rescreened after 30 days from last instance of laceration self-harm in consultation with the LEDP therapist. Participants must have no new psychiatric medications or change in dosage for at least one week prior to randomization. Finally, MRI contraindications do not exclude participants from completing other study visits.

To evaluate eligibility criteria regarding orthostatic hypotension, medical charts are reviewed. Orthostatic hypotension is defined as a drop of ≥20 mmHg in systolic blood pressure or a drop of ≥10 mm Hg in diastolic blood pressure when measured shortly after transitioning from lying down to standing. If evidence of orthostasis is present in chart, study staff consult with LEDP medical provider to determine if the participant is at increased fall risk. If participant is determined to be at increased fall risk (e.g., dizziness upon standing) they are excluded. The current target sample size of 102 participants with anorexia nervosa (AN) was based on the recruitment rate of 2 AN participants per month.

##### *Retention*

To enhance retention, contact information for each enrolled subject including email, home/cell phone, and permanent address is collected at baseline visits. Contact information of a parent or guardian is also collected for participants <18 years.

Additionally, participants are compensated for their time via the standard Laureate Institute for Brain Research (LIBR) rate for survey/behavioral data collection ($20) and scanning time and MRI prep ($50). It is estimated that participants engage in approximately 22.5 hours of survey/behavioral data collection or interventions. Approximately 3 hours of scanning time is compensated for participants, for a total possible compensation of $600 per participant for 25.5 patient hours. Compensation is prorated per half hour for participants who do not complete all aspects of the protocol. Payments are provided to subjects via the ClinCard system (https://clincard.com), a global electronic payment technology (similar to a debit card). After each visit, subjects have their account credited with the amount paid for their participation.

##### *Lost to Follow-Up*

A participant is considered lost to follow-up if they fail to return for a scheduled follow-up visit and study staff are unable to contact the participant after at least three attempts. The following actions must be taken if a participant fails to return for a required study visit: (1) The site will attempt to contact the participant, ascertain if the participant wishes to and/or should continue in the study, and as needed, reschedule the missed visit (within one week of the originally scheduled date). (2) Before a participant is deemed lost to follow-up, the investigator will make every effort to regain contact with the participant (where possible, three telephone calls, three text messages, and, if necessary, an email). These contact attempts are documented in the participant's study file. (3) If there is any concern about the safety of a participant lost to follow-up (e.g., due to previously recorded suicidal ideation) and study staff are unable to reach the individual, then confidentiality may be broken, and the Community Outreach Psychiatric Emergency Services (COPES) hotline (918-744-4800) may be called to help ensure safety of the participant. (4) Should the participant continue to be unreachable, they are considered to have withdrawn from the study with a primary reason of lost to follow-up.

#### Blinding and Randomization:

Participants are be randomized to either the interoceptively focused therapy (IFT) or exteroceptively focused therapy (EFT) condition via a pre-determined randomization sequence provided to the study team by the LIBR Data Management Services (DMS) Core. This is entered blindly by DMS core into the randomization function in Research Electronic Data Capture (REDCap; Harris et al., 2009) at a 1:1 allocation ratio for either condition, stratified by Adolescent or Adult LEDP Unit.

For the aperture task (Keizer et al., 2013; Irvine et al., 2019), participants are randomized using a predetermined randomization order in an Excel spreadsheet. As there are four different task conditions (i.e., the doors starting from open or closed, and estimating one's body or ball passing through the doors), participants are randomized at baseline to determine the order of task conditions. Additionally, participants are counterbalanced for the Somatomap3D task (Ralph-Nearman et al., 2019; Arevian et al., 2020) based on if they complete the ideal or current condition first. During the adjective tasks, the illness-related word ratings are also counter balanced with half of the word appearing at baseline MRI and the other half appearing at follow-up MRI.

#### Intervention:

##### *Floatation-REST*

The Float Clinic and Research Center (FCRC) was constructed at the Laureate Institute for Brain Research (LIBR) between 2014-2015. The laboratory aims to investigate the neurophysiological effects of Reduced Environmental Stimulation Therapy (REST) via floatation, as well as explore its potential as a non-pharmacological therapeutic intervention. Participants float in one of two circular fiberglass pools (Figure 3) that were custom designed for research purposes by Floataway, a British company that has been engineering and manufacturing float pools for the past three decades. The open pool has no enclosure, so the room around the pool was specially constructed to provide a highly reliable and well-calibrated float experience.

The closed pool has a tall enclosure around the pool which participants step into through double glass doors.

Waterproof slip-resistant flooring lines both entire float rooms, and an open shower is located immediately adjacent to the pools. Silent heaters placed under the pool maintain the water at a constant temperature and a dedicated heating, ventilation, and air conditioning system maintains the air at a constant temperature, both of which can be adjusted by the experimenter who remains nearby in a control room. An intercom system allows the participant to freely communicate with the experimenter throughout the float session should any issues arise, and specialized speakers placed around the perimeter of the pools allow the experimenter to communicate with the participant and play music to signal the start and end of the session. There are no cameras, and the outside door is automatically locked to ensure complete privacy throughout the float session. While floating, a blue light-emitting diode (LED) light remains illuminated in the background and can be turned off by the participant using an air switch. Once triggered, the air switch activates an infrared wave detection system so that participants can remain floating and still turn the blue light on and off simply by waving their hand in the air. An additional sensor was placed slightly above the water line to discern when the float pool is occupied. Both the occupy sensor and air switch are linked to digital clocks in the control room, allowing for the automated calculation of the total amount of time that a participant is floating and the total amount of time that a participant is floating with the lights off.

**Exteroceptive attenuation during Floatation-REST.** The pool is filled with reverse osmosis water saturated with ~1,800 pounds of United States Pharmacopoeia grade Epsom salt (magnesium sulfate), creating a salt solution maintained at a specific gravity of ~1.27 which allows for effortless floatation. Visual stimulation is minimized by having an entry door and gasket system which expunges all sources of outside light. In addition, there are no windows inside the float room, and the adjacent room contains a private bathroom that also has no windows, and no lights (which are automatically shut off during the float itself). Thus, when the entry door to the float room is sealed and the blue LED light inside the pool is turned off, the float room is completely dark.

Auditory stimulation is minimized by constructing the float room using multiple layers of sound dampening walls with thick insulation and added soundproofing material, restricting most outside airborne sound from entering the room. Structural sounds transmitted via vibrations in the floor are absorbed by having the pool rest on a bed of 48 butyl rubber springs, effectively isolating the pool from the structure of the building, and preventing all structure-borne noises from entering the water. Gustatory stimulation is minimized by having participants eat at minimum one hour prior to the float and refrain from eating and drinking during the float.

Olfactory stimulation is minimized by tightly controlling any sources of odor. Only unscented cleaning products are used inside the float room, and the participant is instructed to shower beforehand to remove any body odor. In addition, the water disinfection system does not emit any odors during the oxidative process (a major advantage over chlorine).

Beyond the natural sanitation created by having such a high concentration of salt, the water inside the float pool is also circulated through a powerful disinfection system that includes surface skimming, 10-micron filtration, 6 ultraviolet (UV) lights, and 35% Hydrogen Peroxide maintained at ~50ppm. The unique combination of UV with Hydrogen Peroxide generates a free hydroxyl radical that is nearly twice as powerful as chlorine at oxidizing and destroying microorganisms, while also ensuring that no dangerous byproducts or scented odors are emitted during the oxidative process. Between every participant, the water is circulated through this disinfection system a minimum of four full turnovers, providing thorough disinfection of the water.

Thermal stimulation is minimized by calibrating the temperature of the water and air to closely match the temperature of the skin, which is typically a few degrees cooler than core body temperature. Throughout each float session, the water temperature is maintained at ~95.0°F and the air temperature at the rim of the pool is maintained at ~93.5 °F, slightly lower than the water temperature based on the relative humidity of the air in the room. At this temperature setting, it is difficult to discern the boundary between air, body, and water, a unique feature of the float experience. Finally, stimulation from tactile, vestibular and proprioceptive channels is minimized, as is most movement and speech, due to the dense saline solution buffering the body against the forces of gravity and allowing the individual to effortlessly float on their back while remaining completely still (the importance of “stillness” is emphasized in the pre-float instruction set).

***Compliance and Discontinuation:***

Participants are informed that participation is completely voluntary and they are allowed to withdraw from the study at any time. If a participant indicates they are unwilling to do the primary aspects of the study (e.g., floatation session), they are excluded from participation. However, if a subject discontinues from the floatation session after having started a session but does not withdraw from the study, they are allowed to complete the remainder of the study. At the time of study intervention discontinuation or study withdrawal, the reason(s) for discontinuing the participant from the intervention, and methods for determining the need to discontinue would be documented. If a participant withdraws, they are asked to complete the Withdrawn Questionnaire to indicate reasoning for withdrawing. An investigator may discontinue a participant from the study for the following reasons: (1) Significant study intervention non-compliance (i.e., unwilling to complete Floatation-REST sessions or other study related components). (2) Lost-to-follow up; unable to contact subject (see Recruitment and Retention for further information). (3) Any event or medical condition or situation occurs such that continued collection of follow-up study data would not be in the best interest of the participant or might require an additional treatment that would confound the interpretation of the study. (4) The participant meets an exclusion criterion (either newly developed or not previously recognized) that precludes further study participation. To be eligible for follow-ups participants must complete at least one experimental session. If a patient withdraws from the study prior to completing one experimental session they will not be included in data analysis.

**Data Collection**

The collection and processing of personal data from subjects enrolled in this study is limited to those data that are necessary to fulfill the objectives of the study. Only the principal investigator (PI) and trained study personnel have access to linking information or data. The PI is responsible for ensuring the accuracy, completeness, legibility, and timeliness of the data reported. Clinical data (including adverse events (AEs), concomitant medications, mental health diagnoses, and expected adverse reactions data) is entered into the REDCap electronic data capture system provided by LIBR.

***Measures***

See Table 1 for a full list of measures. Additionally, physiological, interview-based, and neuroimaging measures are briefly described below.

**Physiological Measurements**. Anthropomorphic body measurements are taken for comparison with perceptual estimations of body size collected in the string task (Schneider et al., 2009), aperture task (Keizer et al., 2013; Irvine et al., 2019), and Somatomap3D (Ralph-Nearman et al., 2019, 2021; Arevian et al., 2020). Additionally, an InBody is taken to determine participant body composition metrics. Participants are shielded from seeing their body measurements (they are taken from behind the individual to minimize a potential biasing of their estimations) and body composition (screen of InBody is covered to obscure view of weight and body fat percentage) measurements.

**Interview-Based Measures.** At baseline, a brief pre-treatment questionnaire is administered to determine the participants previous exposure to acceptance- and mindfulness-based interventions and Floatation-REST. At the post-intervention session and the 6-month follow up session, participants complete a Feedback Interview. During this, participants are asked about their thoughts and experiences related to the study. The Interviews are tailored to each condition, and queries about if the participant has used or will use any acceptance and mindfulness-based skills introduced in the study in their daily lives.

**Self-Report Measures.** Further self-report measures (besides those measuring BID and interoception) are collected for the exploratory aim to determine possible moderators of the change in perceptual BID. Measures assess: eating disorder symptoms via the Eating Disorder Examination Questionnaire (EDE-Q6; Fairburn, 2008) and Eating Disorder Inventory (EDI, only at 6 month follow up; Garner, 1991), anxiety via the State Trait Anxiety Inventory (STAI; Spielberger et al., 1970) and Anxiety Sensitivity Inventory (ASI; Taylor et al., 2007), body image related to cognitive flexibility via the Body Image Acceptance and Action Questionnaire (BIAAQ-II; Sandoz et al., 2013), affect on the Positive and Negative Affect Schedule (PANAS-X; Watson et al., 1988), depression symptoms via the Patient Health Questionnaire (PHQ-9; Kroenke et al., 2001), sensory sensitivity via the Glasgow Sensory Questionnaire (GSQ; Robertson and Simmons, 2013), avoid/restrictive food intake disorder symptoms via the nine-item Avoidant/Restrictive Food Intake Disorder Screen (ARFID screen; Zickgraf and Ellis, 2018), treatment credibility and expectancies via the Credibility/Expectancies Questionnaire (CEQ; Devilly and Borkovec, 2000), therapeutic alliance via the Working Alliance Inventory – Self Report (WAI-SR; Horvath and Greenberg, 1989), and gastro-intestinal specific anxiety via the Visceral Sensitivity Index (VSI; Labus et al., 2004). Measures of body dissolution and altered states of consciousness via 5 Dimension Altered States of Consciousness Scale (5D-ASC; Studerus et al., 2010) and Body Boundary Dissolution Scale (BBDS; Dambrun, 2016) is explored during experimental sessions. Before and after neuroimaging and experimental sessions, the Karolinska Sleepiness Scale (KSS; Åkerstedt and Gillberg, 1990) is used to determine relative alertness. Prior to scanning, participants complete the Participant Last Use Summary (PLUS; Lisdahl et al., 2018), a list of questions regarding last use of substances including alcohol, marijuana, caffeine, tobacco and other illicit substances. After scanning, participants complete anxiety ratings for illness-related words, as well as valence/arousal ratings for illness-related words. Finally, if a participant withdraws, they are asked to complete a Withdrawn Questionnaire inquiring about reasons for withdrawal.

**Neuroimaging Session.** Prior to entry in the scanner, participants complete an MRI Environment Screen to report any possible metal in the body, and eligibility is confirmed in coordination with LIBR MRI technicians. Additionally, the breathalyzer, and urine drug and pregnancy tests are completed. The subject is then asked to remove all ferromagnetic items. If the subject requires vision correction, they are fitted with plastic glasses closely matching her optic prescription. The audio system is explained so the participant knows how to communicate with the scanner operator. Earplugs are inserted into participants ears, and headphones are placed over the ears. The subject is reminded of the importance of staying still in the scanner. To minimize motion, the participant's head is stabilized with a specially designed pillow under the neck and wedge-shaped cushions between the sides of the head and head-holder. Padding is arranged to maximize comfort, and a pillow is placed under the knees to provide lower back relief. The MRI technician places the head coil in position over the participant's head, localizes the head position on the scanner, and ensures that the subject can fully view the display screen at the end of the scanner gurney, where task images are displayed, by looking in the mirror directly above their eyes.

During the baseline behavioral session, participants complete the Somatomap3D task with an MRI-compatible trackball to gain practice using the apparatus. During the MRI session, the MRI-compatible trackball is positioned in the subject's dominant hand and the subject tests the buttons. In the subject's non-dominant hand, the pulse oximeter is placed on one finger, and the subject is provided the emergency squeeze ball. Once the subject is comfortable, the gurney moves into the magnet. The MRI technician secures the scan room door, dims the scan room lights, and communicates with the participant through the console.

During scanning, participants complete tasks including a clinical scan, structural T1, resting state scan (to measure brain network activity at rest), Visceral Interoceptive Attention (VIA; Simmons et al., 2013) task, a modified version of the Adjective Task (Matthews et al., 2010), and the Somatomap3D (Ralph-Nearman et al., 2019, 2021; Arevian et al., 2020). During the resting state scan, participants are asked to keep their head as still as possible and to keep their eyes open and fixated on a white cross presented at the center of a dark screen, and to "clear your mind and do not think about anything in particular". The duration of resting state scanning is about 24 minutes. Participants additionally complete a T1-weighted structural scan of the brain is obtained. Time to complete this scan is about six minutes.

The modified version of the Adjective Task (Matthews et al., 2010) probes self-referential processing using three types of trials: self-evaluation, other evaluation, and word evaluation. An adjective is presented in each trial, and participants are prompted to evaluate whether that adjective describes him/herself (self-evaluation, e.g. "I am…?"), another person (other evaluation, e.g. "My best friend is…?"), or whether the word has positive valence (word evaluation, e.g. "The word is positive…?"). In each case, participants use a trackball to indicate yes/no responses. The task has been modified to include illness-related adjectives (e.g., skinny, fat, hungry, bloated). Following the scanning, participants are asked to rate illness-related words from the task for first anxiety, and then valence/arousal levels. Two different sets of words are counterbalanced across participants to account for potential test-retest issues from baseline to post-intervention. Scanning procedures take place at two time points (baseline and post-intervention) for the current study and scanning lasts about 90 minutes each.

**Design Considerations**

The following considerations were taken into account in the design of the current protocol:

(1) Why are we not utilizing water loading tasks or other gastric interoception tasks? While gastric interoception may be more closely related to AN psychophysiology, historically

there have been challenges in examining gastrointestinal perception including the invasiveness of certain approaches (e.g., inflatable balloons) and inability to repeatedly engage in water loading procedures (Khalsa et al., 2022). Novel minimally invasive probes are available (Smith et al., 2021b); however, they are still being validated by our group at this time (R01MH127225). Future studies should include measures of gastrointestinal interoception.

(2) Will engaging in cognitive exercises be feasible during Floatation-REST? Engaging in cognitive exercises while floating may detract from the interoceptive manipulation. Thus, minimal instruction will be given to participants in this condition. Instead, their experience will be assessed in the post-floatation surveys. We still expect to see a reduction in BID in both the IFT and EFT groups.

(3) Why were these measures of BID chosen? There are other ways to measure both cognitive/affective and perceptual BID that were considered. A broad range of measures have been chosen to capture BID based on previous studies conducted with Floatation-REST in AN patients (i.e., PFRS; Swami et al., 2008) and cognitive/affective measures we expect to be targeted via mindfulness and acceptance skills (i.e., BISS; Cash et al., 2002—a state measure, and MBSRQ; Cash, 2000, specifically the appearance scales).

(4) What if manipulating interoception via Floatation-REST does not reduce perceptual BID? This is the purpose of the exploratory analyses to determine what is the target of a possible experimental therapeutic approach for the reduction of perceptual BID. The measurement of other possible mechanisms will allow for the identification of possible targets for future study in the development of such an approach. Other manipulations of interoceptive via heartbeat perception (Heartbeat Tapping Task; Ludwick-Rosenthal and Neufeld, 1985; Canales-Johnson et al., 2015; Smith et al., 2021a) and respiratory perception (Inspiratory Breath Hold Task; Lapidus et al., 2020) are being investigated.

(5) How can we minimize impacts of biological variation? We only recruit participants with a BMI ≥ 17.5 to avoid potential transient impacts of acute starvation on cognition and interoception.

**Analyses**

***Secondary Analyses***

Hypotheses 2a-c: To test the hypothesis that measures of perceptual and cognitive/affective BID will relate to interoception:

1) Correlations among measures of perceptual—including behavioral (Schneider et al., 2009; Keizer et al., 2013; Irvine et al., 2019) and self-report (Swami et al., 2008; Ralph-Nearman et al., 2021)—and cognitive/affective BID (self-report; Cash, 2000), as well as interoception—self-report (Mehling et al., 2012; Gabriele et al., 2022) and behavioral (Lapidus et al., 2020; Smith et al., 2020)—will be examined.
2) Linear regression models will be fit to predict self-reported and behavioral measures of BID (both perceptual and cognitive/affective) from self-report and behavioral assays of interoception assessed at baseline.
3) Finally, to examine the strength of cross-sectional relationships between BID and interoception, Fisher's r to z transformation will be conducted so that z-score comparisons can determine significant differences among correlations.

***Exploratory Analyses***

Hypothesis 3a: Group differences in perceptual BID (Swami et al., 2008; Ralph-Nearman et al., 2019, 2021) will be explored at six-week and six-month follow-up using LMM. In addition to group, baseline and post-intervention interoception, cognitive/affective BID, and eating disorder symptomatology will be entered as regressors of interest.

Hypothesis 3b: Multilevel mediation models will be fit to explore perceptual BID changes via multimodal interoception, and state/trait illness measures of AN. Potential mediators include eating disorder symptoms, state anxiety during experimental session, and interoception during experimental session. Individual differences in baseline interoception will be examined as a mediator of perceptual BID outcomes. Further, this exploratory mediation analysis will investigate if there is a moderating effect of baseline cognitive/affective BID on perceptual BID change.

Hypothesis 3c: Analyses will be conducted to examine changes on self-report measures, as well as brain activation and connectivity.

**Safety**

***Data and Safety Monitoring and Reporting***

While the procedures involved in the proposed protocol represent minimal added risk for adverse events, the study population (females with AN), are at increased risk for negative events (e.g., suicidal ideation, hypotension, significantly low weight). Data safety and monitoring is the responsibility of the PI (Dr. Emily Choquette, licensed clinical psychologist) and mentorship team (Dr. Sahib Khalsa, board-certified psychiatrist, and Dr. Robin Aupperle, licensed clinical psychologist) in concert with the institutional review board (IRB). Safety monitoring involves review of accumulated outcome data for groups of participants to determine if any of the procedures practiced should be altered or stopped. Drs. Khalsa and Aupperle are available for consultation for any medical or safety issues. Additionally, close monitoring of participant safety includes prompt reporting of adverse events to the IRB.

In addition to safety monitoring, Dr. Choquette (PI) is responsible for complying with the reporting requirements and also works with an independent safety monitor (ISM). A psychiatrist from the Tulsa community, Dr. Matthew Meyer, serves as the ISM on the protocol. The role of the ISM is to provide independent clinical oversight. The ISM receives summaries of all adverse events in real time and annual reports concerning monitoring for adverse medical and psychiatric events. The ISM has access to all assessment data (through REDCap; Harris et al., 2009) and is able to assess the participant at any time during the study in order to monitor clinical risk as well as whether research participation may not be in the participant's best interest.

This study does not provide rescue therapy. In case that participants experience worsening symptoms, their participation may be terminated, and the participants would be referred to their clinician within the LEDP or if worsening is at virtual follow up the participant would be referred to other local clinicians.

Dr. Choquette provides a summary of the safe conduct of the study to NIH as part of the annual progress report, which parallels the written report required by the IRB for annual project renewal. All immediately life-threatening events, whether related or unrelated, will be recorded on the serious adverse event (SAE) Form and submitted to the ISM and LIBR Human Protections and Compliance officer within 24 hours of site awareness. Human Protections and Compliance Officer will help determine if the event is immediately reportable to IRB or can wait until continuing review. Study sponsor will be notified within 48 hours of site awareness. Review of data and procedures may result in early termination of the study, amendments to the protocol, or changes to the data collection plan. Should the protocol or data collection plans be amended as a result of data review, the IRB would be notified, and the amendment approved prior to study amendment implementation. In addition, participants are notified of any significant new risks that develop during the course of research that may affect their wish to continue study participation.

***Monitoring for Medical Risk and Relapse***

Blood pressure is monitored via medical chart and taken up to three times per day on the unit. Further, study staff and LEDP providers communicate should there be any medical changes or concerns. There is little consensus on the definition of relapse in the field; as such, we take a medical monitoring approach. The majority of participants are in residential care through the post-intervention time point, and therefore they are carefully monitored from a relapse-prevention standpoint by their LEDP provider.

***Monitoring for Suicidal Ideation***

The Patient Health Questionnaire (PHQ-9; Kroenke et al., 2001) is administered at baseline, post-intervention and remote follow up sessions. Endorsement of item 9 during any of these time points triggers a safety assessment. Should a patient indicate intent or plan during this assessment, the patient will be referred to the LEDP treatment team or to a local medical provider in their community (or the 911/988 hotlines if there is cause of immediate concern) for additional care, and the ISM will be notified to determine if the patient should be withdrawn from the study. The Columbia-Suicide Severity Rating Scale (C-SSRS; Posner, 2016) is additionally completed at baseline and post-intervention for all participants. Participants who indicate any ideation greater than passive infrequent ideation would complete a safety plan with a trained study staff member if they do not have one already at LEDP. The safety plan would be given to the participant as well as information regarding who to contact if their ideation worsens or they develop a plan or intent.

#### Additional SPIRIT Checklist Items

**Spirit Item 2**

**Supplementary Table 1**

*Protocol Version and Amendments*

| Version | Date | Description of Change | Brief Rationale |
|---|---|---|---|
| V 2.0 | 3/26/2024 | 1. Modification of intervention description<br>2. Additional measures or | Modifications were made to finalize protocol prior to study start. Changes made through |

| | | | |
|---|---|---|---|
| | | modification to measures<br>3. Updated protocol and consent to reflect changes made to protocol, update to funding and PI status<br>4. Informed consent modified to include NDA information required by NIH funding status | natural development of intervention. Additional measures and updated measures to better capture constructs of interest. Additional changes were made to reflect changing status of the grant as funded and PI's status. |
| V 3.0 | 9/16/2024 | 1. Additional measures<br>2. Additional safety monitoring<br>3. Change in BMI criteria from 18.5 to 17.5<br>4. Flexibility of baseline and post-intervention sessions<br>5. Update NCT number and contact phone number for PI | Modifications were made to include additional constructs of interest, address the reporting of adverse events (unrelated to study), and to enhance ability to recruit participants with enough time to complete the study before discharging from LEDP. |
| V 3.1 | 1/3/2025 | 1. Update to protocol regarding safety reporting | Modifications made to describe suicidal ideation as an expected event, given the study population and with monitoring at each timepoint. |
| V 4.0 | 3/21/2025 | 1. Age range changed from 15-40 to 13-50 | Modifications were made to expand inclusion of potential participants and investigate outcomes across a longer age-range. |
| V 5.0 (current protocol) | 9/24/2025 | 1. Change in orthostatic hypotension criteria to include LEDP provider consult to ascertain if participants are at a clinical fall risk | Modifications were made to enhance recruitment ability while prioritizing participant safety. |

*Note.* The table above is intended to capture changes of IRB-approved versions of the protocol, including a description of the change and rationale.

**Spirit Item 3d**

The single coordinating site in this study is the LIBR, where LIBR study staff are the steering committee involved in data collection and management in concert with the IRB. Listed below are individuals of the study team and their respective roles.

***Principal Investigator***

Dr. Emily Choquette (EMC), Ph.D., licensed clinical psychologist, serves as the principal investigator (PI) on the current study. Dr. Choquette is Associate Investigator at LIBR and Assistant Research Professor at the University of Tulsa. As the PI, Dr. Choquette is responsible for leading the current study safely and ethically, as well as training research staff to manage day-to-day trial coordination.

***K23 Award Mentors and Consultants***

Mentors include Drs. Sahib Khalsa, M.D., Ph.D., Robin Aupperle, Ph.D., Dr. Nancy Zucker, Ph.D. The primary mentor for the study is Dr. Khalsa, Associate Professor in Residence in the Psychiatry Department at the University of California, Los Angeles Geffen School of Medicine, Director of Anxiety Disorders Research at the University of California, Los Angeles Semel Institute for Neuroscience and Human Behavior, and Affiliate Investigator at LIBR. Dr. Khalsa's research examines the role of interoception in psychiatric disorders using pharmacological and nonpharmacological methodologies and investigates potential therapeutic applications of interoceptive interventions via clinical trials. As primary mentor and board-certified psychiatrist, Dr. Khalsa provides oversight of the study aid with scientific direction in the execution of the research plan.

A co-mentor for this study is Dr. Aupperle, LIBR Principal Investigator, Director of Clinical Practicum at LIBR, as well as Associate Professor at University of Tulsa. Dr. Aupperle is a licensed clinical psychologist, whose research specializes in brain-behavior relationships, and the development of neuroscience informed behavioral interventions for depression, anxiety, and common co-occurring symptoms/conditions. She provides mentorship to the principal investigator in the day-to-day execution of research activities. Additionally, a co-mentor for the study is Dr. Zucker, Professor in the Department of Psychiatry and Behavioral Sciences at Duke University Medical Center and Department of Psychology and Neuroscience at Duke University, and Founder and Director of the Duke Center for Eating Disorders. Dr. Zucker's research focuses on examining the role of interoceptive dysfunction in eating and feeding disorders. She provides mentorship to the principal investigator focused on neurobiology of eating disorders, as well as general guidance on the execution of the research.

Consultants include Drs. Scott Moseman, M.D., Karen Quigley, Ph.D., and Wes Thompson, Ph.D. Dr. Moseman is a board-certified psychiatrist and Medical Director of the LEDP, who will serve as consultant on the study regarding clinical and safety-related monitoring.

Dr. Quigley is Professor of Psychology in the Northeastern University College of Sciences, and an expert in psychophysiological measurement of interoception and will consult with the principal investigator on topics related to physiological measurement (i.e., heart rate, blood pressure, and respiration), processing of physiological data, and statistical methodology. Dr. Wesley Thompson is Principal Investigator of the Center for Population Neuroscience and Genetics at LIBR and an adjunct professor at the Herbert Wertheim School of Public Health & Human Longevity Science, at the University of California, San Diego. He has experience in Bayesian modeling and longitudinal data analysis and will serve as the expert statistical consultant for the current study.

Dr. Matthew Meyer, M.D., is a board-certified psychiatrist from the Tulsa area who serves as Independent Safety Monitor (ISM). Dr Meyer provides clinical insight on the current study and has extensive prior ISM experience with LIBR studies.

**Spirit Item 11**

There was no patient or public involvement in the design of this trial.

#### References

Åkerstedt, T., and Gillberg, M. (1990). Subjective and objective sleepiness in the active individual. *Int. J. Neurosci.* 52, 29–37. doi: 10.3109/00207459008994241

Arevian, A. C., O'Hora, J., Rosser, J., Mango, J. D., Miklowitz, D. J., and Wells, K. B. (2020). Patient and provider cocreation of mobile texting apps to support behavioral health: usability study. *JMIR mHealth uHealth* 8, e12655. doi: 10.2196/12655

Canales-Johnson, A., Silva, C., Huepe, D., Rivera-Rei, Á., Noreika, V., Garcia, M. del C., et al. (2015). Auditory feedback differentially modulates behavioral and neural markers of objective and subjective performance when tapping to your heartbeat. *Cereb. Cortex* 25, 4490–4503. doi: 10.1093/cercor/bhv076

Cash, T. F. (2000). The Multidimensional Body-Self Relations Questionnaire Users' Manual. Norfolk, VA: Old Dominion University.

Cash, T. F., Fleming, E. C., Alindogan, J., Steadman, L., and Whitehead, A. (2002). Beyond body image as a trait: the development and validation of the Body Image States Scale. *Eat. Disord.* 10, 103–113. doi: 10.1080/10640260290081678

Dambrun, M. (2016). When the dissolution of perceived body boundaries elicits happiness: the effect of selflessness induced by a body scan meditation. *Conscious. Cogn.* 46, 89–98. doi: 10.1016/j.concog.2016.09.013

Devilly, G. J., and Borkovec, T. D. (2000). Psychometric properties of the credibility/expectancy questionnaire. *J. Behav. Ther. Exp. Psychiatry* 31, 73–86. doi: 10.1016/s0005-7916(00)00012-4

Fairburn, C. G. (2008). Cognitive Behavior Therapy and Eating Disorders. New York: Guilford Press.

Gabriele, E., Spooner, R., Brewer, R., and Murphy, J. (2022). Dissociations between self-reported interoceptive accuracy and attention: evidence from the Interoceptive Attention Scale. *Biol. Psychol.* 168, 108243. doi: 10.1016/j.biopsycho.2021.108243

Garner, D. M. (1991). Eating Disorder Inventory-2; Professional Manual. Odessa, FL: Psychological Assessment Resources.

Harris, P. A., Taylor, R., Thielke, R., Payne, J., Gonzalez, N., and Conde, J. G. (2009). Research electronic data capture (REDCap)—a metadata-driven methodology and workflow process for providing translational research informatics support. *J. Biomed. Inform.* 42, 377–381. doi: 10.1016/j.jbi.2008.08.010

Horvath, A. O., and Greenberg, L. S. (1989). Development and validation of the Working Alliance Inventory. *J. Couns. Psychol.* 36, 223–233. doi: 10.1037/0022-0167.36.2.223

Irvine, K. R., McCarty, K., McKenzie, K. J., Pollet, T. V., Cornelissen, K. K., Tovée, M. J., et al. (2019). Distorted body image influences body schema in individuals with negative bodily attitudes. *Neuropsychologia* 122, 38–50. doi: 10.1016/j.neuropsychologia.2018.11.015

Keizer, A., Smeets, M. A. M., Dijkerman, H. C., Uzunbajakau, S. A., van Elburg, A., and Postma, A. (2013). Too fat to fit through the door: first evidence for disturbed body-scaled action in anorexia nervosa during locomotion. *PLoS One* 8, e64602. doi: 10.1371/journal.pone.0064602

Khalsa, S. S., Berner, L. A., and Anderson, L. M. (2022). Gastrointestinal interoception in eating disorders: charting a new path. *Curr. Psychiatry Rep.* 24, 47–60. doi: 10.1007/s11920-022-01318-3

Kroenke, K., Spitzer, R. L., and Williams, J. B. (2001). The PHQ-9: validity of a brief depression severity measure. *J. Gen. Intern. Med.* 16, 606–613. doi: 10.1046/j.1525-1497.2001.016009606.x

Labus, J. S., Bolus, R., Chang, L., Wiklund, I., Naesdal, J., Mayer, E. A., et al. (2004). The Visceral Sensitivity Index: development and validation of a gastrointestinal symptom-specific anxiety scale. *Aliment. Pharmacol. Ther.* 20, 89–97. doi: 10.1111/j.1365-2036.2004.02007.x

Lapidus, R. C., Puhl, M., Kuplicki, R., Stewart, J. L., Paulus, M. P., Rhudy, J. L., et al. (2020). Heightened affective response to perturbation of respiratory but not pain signals in eating, mood, and anxiety disorders. *PLoS One* 15, e0235346. doi: 10.1371/journal.pone.0235346

Lisdahl, K. M., Sher, K. J., Conway, K. P., Gonzalez, R., Feldstein Ewing, S. W., Nixon, S. J., et al. (2018). Adolescent brain cognitive development (ABCD) study: overview of substance use assessment methods. *Dev. Cogn. Neurosci.* 32, 80–96. doi: 10.1016/j.dcn.2018.02.007

Ludwick-Rosenthal, R., and Neufeld, R. W. (1985). Heart beat interoception: a study of individual differences. *Int. J. Psychophysiol.* 3, 57–65. doi: 10.1016/0167-8760(85)90020-0

Matthews, S. C., Simmons, A. N., Strigo, I. A., Arce, E., Stein, M. B., and Paulus, M. P. (2010). Escitalopram attenuates posterior cingulate activity during self-evaluation in healthy volunteers. *Psychiatry Res.* 182, 81–87. doi: 10.1016/j.pscychresns.2010.02.003

Mehling, W. E., Price, C., Daubenmier, J. J., Acree, M., Bartmess, E., and Stewart, A. (2012). The Multidimensional Assessment of Interoceptive Awareness (MAIA). *PLoS One* 7, e48230. doi: 10.1371/journal.pone.0048230

Posner, K. (2016). Columbia-Suicide Severity Rating Scale. APA PsycTests. doi: 10.1037/t52667-000

Ralph-Nearman, C., Arevian, A. C., Moseman, S., Sinik, M., Chappelle, S., Feusner, J. D., et al. (2021). Visual mapping of body image disturbance in anorexia nervosa reveals objective markers of illness severity. *Sci. Rep.* 11, 12262. doi: 10.1038/s41598-021-90739-w

Ralph-Nearman, C., Arevian, A. C., Puhl, M., Kumar, R., Villaroman, D., Suthana, N., et al. (2019). A novel mobile tool (Somatomap) to assess body image perception pilot tested with fashion models and nonmodels: cross-sectional study. *JMIR Ment. Health* 6, e14115. doi: 10.2196/14115

Robertson, A. E., and Simmons, D. R. (2013). The relationship between sensory sensitivity and autistic traits in the general population. *J. Autism Dev. Disord.* 43, 775–784. doi: 10.1007/s10803-012-1608-7

Sandoz, E. K., Wilson, K. G., Merwin, R. M., and Kellum, K. (2013). Assessment of body image flexibility: the Body Image-Acceptance and Action Questionnaire. *J. Contextual Behav. Sci.* 2, 39–48. doi: 10.1016/j.jcbs.2013.03.002

Schneider, N., Frieler, K., Pfeiffer, E., Lehmkuhl, U., and Salbach-Andrae, H. (2009). Comparison of body size estimation in adolescents with different types of eating disorders. *Eur. Eat. Disord. Rev.* 17, 468–475. doi: 10.1002/erv.956

Simmons, W. K., Avery, J. A., Barcalow, J. C., Bodurka, J., Drevets, W. C., and Bellgowan, P. (2013). Keeping the body in mind: insula functional organization and functional connectivity integrate interoceptive, exteroceptive, and emotional awareness. *Hum. Brain Mapp.* 34, 2944–2958. doi: 10.1002/hbm.22113

Smith, R., Feinstein, J. S., Kuplicki, R., Forthman, K. L., Stewart, J. L., Paulus, M. P., et al. (2021a). Perceptual insensitivity to the modulation of interoceptive signals in depression, anxiety, and substance use disorders. *Sci. Rep.* 11, 2108. doi: 10.1038/s41598-021-81307-3

Smith, R., Kuplicki, R., Feinstein, J., Forthman, K. L., Stewart, J. L., Paulus, M. P., et al. (2020). A Bayesian computational model reveals a failure to adapt interoceptive precision estimates across depression, anxiety, eating, and substance use disorders. *PLoS Comput. Biol.* 16, e1008484. doi: 10.1371/journal.pcbi.1008484

Smith, R., Mayeli, A., Taylor, S., Al Zoubi, O., Naegele, J., and Khalsa, S. S. (2021b). Gut inference: a computational modelling approach. *Biol. Psychol.* 164, 108152. doi: 10.1016/j.biopsycho.2021.108152

Spielberger, C., Gorsuch, R., and Lushene, R. (1970). Manual for the State-Trait Anxiety Inventory. Palo Alto, CA: Consulting Psychologists Press.

Studerus, E., Gamma, A., and Vollenweider, F. X. (2010). Psychometric evaluation of the Altered States of Consciousness Rating Scale (OAV). *PLoS One* 5, e12412. doi: 10.1371/journal.pone.0012412

Swami, V., Salem, N., Furnham, A., and Tovée, M. J. (2008). Initial examination of the validity and reliability of the female photographic figure rating scale for body image assessment. *Pers. Individ. Dif.* 44, 1752–1761. doi: 10.1016/j.paid.2008.02.002

Taylor, S., Zvolensky, M. J., Cox, B. J., Deacon, B., Heimberg, R. G., Ledley, D. R., et al. (2007). Robust dimensions of anxiety sensitivity: development and initial validation of the Anxiety Sensitivity Index-3. *Psychol. Assess.* 19, 176–188. doi: 10.1037/1040-3590.19.2.176

Watson, D., Clark, L. A., and Tellegen, A. (1988). Development and validation of brief measures of positive and negative affect: the PANAS scales. *J. Pers. Soc. Psychol.* 54, 1063–1070. doi: 10.1037//0022-3514.54.6.1063

Zickgraf, H. F., and Ellis, J. M. (2018). Initial validation of the Nine Item Avoidant/Restrictive Food Intake disorder screen (NIAS): a measure of three restrictive eating patterns. *Appetite* 123, 32–42. doi: 10.1016/j.appet.2017.11.111
