## Supplementary C for "Interoceptive mechanisms of body image disturbance in anorexia nervosa: Study protocol of a randomized parallel-group trial"

### Certificate of Action

| | |
|---|---|
| **Investigator Name:** Emily M Choquette, PhD | **Board Action Date:** 09/26/2023 |
| **Investigator Address:** 6655 S Yale Ave<br>Tulsa, OK 74136, United States | **Approval Expires:** 09/26/2024<br>**Continuing Review Frequency:** Annually |
| **Sponsor:** Laureate Institute for Brain Research<br>**Institution Tracking Number:** 2023-007 | **Sponsor Protocol Number:** 2023-007<br>**Amended Sponsor Protocol Number:** |
| **Study Number:** 1361152 | **IRB Tracking Number:** 20234215 |
| **Work Order Number:** 1-1697264-1 | |
| **Protocol Title:** Interoceptive mechanisms of body image disturbance in anorexia nervosa | |

**THE FOLLOWING ITEMS ARE APPROVED:**
Investigator
Demographics And Psychosocial #38519613.0 - As Submitted (source: 2023 007_demographicsandpsychosocial)
Medical History #38519614.0 - As Submitted (source: 2023 007_medicalhistory)
Medical History Documentation #38519611.0 - As Submitted (source: 2023 007_medicalhistorydocumentation)
Post-intervention feedback interview #38519612.0 - As Submitted (source: 2023 007_qualitative questions final)
Protocol (09-10-2023) v1.0 (source: 2023 007 abc float k32_protocol_09182023)
Consent Form [IN0-0]

**Please note the following information:**
The Board requires that all subjects must be able to consent for themselves to be enrolled in this study. This means that you cannot enroll incapable subjects who require enrollment by consent of a legally authorized representative.

**THE IRB HAS APPROVED THE FOLLOWING LOCATIONS TO BE USED IN THE RESEARCH:**
Laureate Institute for Brain Research, 6655 S Yale Ave, Tulsa, Oklahoma 74136

**ALL IRB APPROVED INVESTIGATORS MUST COMPLY WITH THE FOLLOWING:**
As a requirement of IRB approval, the investigators conducting this research will:

- Comply with all requirements and determinations of the IRB.
- Protect the rights, safety, and welfare of subjects involved in the research.
- Personally conduct or supervise the research.
- Conduct the research in accordance with the relevant current protocol approved by the IRB.
- Ensure that there are adequate resources to carry out the research safely.
- Ensure that research staff are qualified to perform procedures and duties assigned to them during the research.
- Submit proposed modifications to the IRB prior to their implementation.
    - Not make modifications to the research without prior IRB review and approval unless necessary to eliminate apparent immediate hazards to subjects.
- For research subject to continuing review, submit continuing review reports when requested by the IRB.
- Submit a closure form to close research (end the IRB's oversight) when:
    - The protocol is permanently closed to enrollment
    - All subjects have completed all protocol related interventions and interactions
    - For research subject to federal oversight other than FDA:
        - No additional identifiable private information about the subjects is being obtained
        - Analysis of private identifiable information is completed
- For research subject to continuing review, if research approval expires, stop all research activities and immediately contact the IRB.

This is to certify that the information contained herein is true and correct as reflected in the records of WCG IRB. WE CERTIFY THAT WCG IRB IS IN FULL COMPLIANCE WITH GOOD CLINICAL PRACTICES AS DEFINED UNDER THE U.S. FOOD AND DRUG ADMINISTRATION (FDA) REGULATIONS, U.S. DEPARTMENT OF HEALTH AND HUMAN SERVICES (HHS) REGULATIONS, AND THE INTERNATIONAL CONFERENCE ON HARMONISATION (ICH) GUIDELINES.

- Promptly (within 5 days) report to the IRB the information items listed in the IRB's "Prompt Reporting Requirements" available on the IRB's Web site.
- Not accept or provide payments to professionals in exchange for referrals of potential subjects ("finder's fees.")
- Not accept payments designed to accelerate recruitment that are tied to the rate or timing of enrollment ("bonus payments") without prior IRB approval.
- When required by the IRB ensure that consent, permission, and assent are obtained and documented in accordance with the relevant current protocol as approved by the IRB.
- Promptly notify the IRB of any change to information provided on your initial submission form.

Consistent with AAHRPP's requirements in connection with its accreditation of IRBs, the individual and/or organization shall promptly communicate or provide, the following information relevant to the protection of human subjects to the IRB in a timely manner:

- Upon request of the IRB, a copy of the written plan between sponsor or CRO and site that addresses whether expenses for medical care incurred by human subject research subjects who experience research related injury will be reimbursed, and if so, who is responsible in order to determine consistency with the language in the consent document.
- Any site monitoring report that directly and materially affects subject safety or their willingness to continue participation. Such reports will be provided to the IRB within 5 days.
- Any findings from a closed research when those findings materially affect the safety and medical care of past subjects. Findings will be reported for 2 years after the closure of the research.

For Investigator's Brochures, an approval action indicates that the IRB has the document on file for the research.

When the Board approves subject materials and/or advertisements, any redline changes that were provided by the submitter or required by the Board for approval will remain visible in the outcome document(s); however, recipients are expected to accept the tracked changes in the document before using. Do not make any additional modifications (including font size and visual effects) to the approved materials.
If the IRB approved an e-consent process that involves uploading the approved consent form to an e-consent platform, please ensure that the consent form(s) approved for your site is the version of the consent form that gets uploaded to the platform.

If the board approves a change of Principal Investigator - Once approved, the new Principal Investigator is authorized by WCG IRB to carry out the study as previously approved for the prior Principal Investigator (unless the Board provides alternate instructions to the new Principal Investigator). This includes continued use of the previously approved study materials. The IRB considers the approval of the new PI a continuation of the original approval, so the identifying information about the study remains the same.

If your research site is a HIPAA covered entity, the HIPAA Privacy Rule requires you to obtain written authorization from each research subject for any use or disclosure of protected health information for research. If your IRB-approved consent form does not include such HIPAA authorization language, the HIPAA Privacy Rule requires you to have each research subject sign a separate authorization agreement.

If this study includes data monitoring committee/data safety monitoring board, please note that the reports of all meetings of this committee should be submitted to the IRB even if the outcome of the meeting results in no changes to the study.

**For research subject to continuing review, you will receive Continuing Review Report forms from WCG IRB when the expiration date is approaching.**

Thank you for using this WCG IRB to provide oversight for your research project.

**DISTRIBUTION OF COPIES:**
**<u>Contact, Company</u>**
Colleen McCallum, MBA, MSW, Laureate Institute for Brain Research
Teresa Victor, Laureate Institute for Brain Research
Emily M Choquette, PhD, Laureate Institute for Brain Research
