## Supplementary D for "Interoceptive mechanisms of body image disturbance in anorexia nervosa: Study protocol of a randomized parallel-group trial"

### Department of Health and Human Services
National Institutes of Health
NATIONAL INSTITUTE OF MENTAL HEALTH

**Notice of Award**
FAIN# K23MH135213
**Federal Award Date**
11/10/2023

#### Recipient Information

**1. Recipient Name**
LAUREATE INSTITUTE FOR BRAIN RESEARCH, LLC
6655 S YALE AVE
TULSA, OK 74136

**2. Congressional District of Recipient**
01

**3. Payment System Identifier (ID)**
1731328881A1

**4. Employer Identification Number (EIN)**
731328881

**5. Data Universal Numbering System (DUNS)**
967230579

**6. Recipient's Unique Entity Identifier**
MX8ULDF6SDD6

**7. Project Director or Principal Investigator**
Emily Choquette, PHD


918-703-2343

**8. Authorized Official**
Colleen McCallum


#### Federal Agency Information

**9. Awarding Agency Contact Information**
Christine Clarkson
Grants Management Specialist
NATIONAL INSTITUTE OF MENTAL HEALTH

301-402-5756

**10. Program Official Contact Information**
Mark Chavez
Health Scientist Administrator
NATIONAL INSTITUTE OF MENTAL HEALTH

301-443-8942

#### Federal Award Information

**11. Award Number**
1K23MH135213-01

**12. Unique Federal Award Identification Number (FAIN)**
K23MH135213

**13. Statutory Authority**
42 USC 241 42 CFR 52

**14. Federal Award Project Title**
Interoceptive mechanisms of body image disturbance in anorexia nervosa

**15. Assistance Listing Number**
93.242

**16. Assistance Listing Program Title**
Mental Health Research Grants

**17. Award Action Type**
New Competing

**18. Is the Award R&D?**
Yes

##### Summary Federal Award Financial Information

| | |
|---|---|
| **19. Budget Period Start Date** 12/01/2023 **– End Date** 11/30/2024 | |
| **20. Total Amount of Federal Funds Obligated by this Action** | $158,035 |
| 20 a. Direct Cost Amount | $146,329 |
| 20 b. Indirect Cost Amount | $11,706 |
| **21.** Authorized Carryover | |
| **22.** Offset | |
| **23.** Total Amount of Federal Funds Obligated this budget period | $158,035 |
| **24. Total Approved Cost Sharing or Matching, where applicable** | $0 |
| **25. Total Federal and Non-Federal Approved this Budget Period** | $158,035 |
| **26. Project Period Start Date** 12/01/2023 **– End Date** 11/30/2028 | |
| **27.** Total Amount of the Federal Award including Approved Cost Sharing or Matching this Project Period | $158,035 |

**28. Authorized Treatment of Program Income**
Additional Costs

**29. Grants Management Officer - Signature**
Theresa A. Mercogliano

**30. Remarks**
Acceptance of this award, including the "Terms and Conditions," is acknowledged by the recipient when funds are drawn down or otherwise requested from the grant payment system.

**SECTION I – AWARD DATA – 1K23MH135213-01**

**Principal Investigator(s):**
Emily Choquette, PHD

Dear Authorized Official:

The National Institutes of Health hereby awards a grant in the amount of $158,035 (see "Award Calculation" in Section I and "Terms and Conditions" in Section III) to LAUREATE INSTITUTE FOR BRAIN RESEARCH in support of the above referenced project. This award is pursuant to the authority of 42 USC 241 42 CFR 52 and is subject to the requirements of this statute and regulation and of other referenced, incorporated or attached terms and conditions.

Acceptance of this award, including the "Terms and Conditions," is acknowledged by the recipient when funds are drawn down or otherwise requested from the grant payment system.

Each publication, press release, or other document about research supported by an NIH award must include an acknowledgment of NIH award support and a disclaimer such as "Research reported in this publication was supported by the National Institute Of Mental Health of the National Institutes of Health under Award Number K23MH135213. The content is solely the responsibility of the authors and does not necessarily represent the official views of the National Institutes of Health." Prior to issuing a press release concerning the outcome of this research, please notify the NIH awarding IC in advance to allow for coordination.

Award recipients must promote objectivity in research by establishing standards that provide a reasonable expectation that the design, conduct and reporting of research funded under NIH awards will be free from bias resulting from an Investigator's Financial Conflict of Interest (FCOI), in accordance with the 2011 revised regulation at 42 CFR Part 50 Subpart F. The Institution shall submit all FCOI reports to the NIH through the eRA Commons FCOI Module. The regulation does not apply to Phase I Small Business Innovative Research (SBIR) and Small Business Technology Transfer (STTR) awards. Consult the NIH website http://grants.nih.gov/grants/policy/coi/ for a link to the regulation and additional important information.

If you have any questions about this award, please direct questions to the Federal Agency contacts.

Sincerely yours,

Theresa A. Mercogliano
Grants Management Officer
NATIONAL INSTITUTE OF MENTAL HEALTH

Additional information follows

**Cumulative Award Calculations for this Budget Period (U.S. Dollars)**

| | |
|---|---|
| **Salaries and Wages** | $77,063 |
| **Fringe Benefits** | $19,266 |
| **Personnel Costs (Subtotal)** | $96,329 |
| **Other** | $50,000 |
| **Federal Direct Costs** | $146,329 |
| **Federal F&A Costs** | $11,706 |
| **Approved Budget** | $158,035 |
| **Total Amount of Federal Funds Authorized (Federal Share)** | $158,035 |
| **TOTAL FEDERAL AWARD AMOUNT** | $158,035 |
| **AMOUNT OF THIS ACTION (FEDERAL SHARE)** | $158,035 |

| **SUMMARY TOTALS FOR ALL YEARS** (for this Document Number) | | |
|---|---|---|
| **YR** | **THIS AWARD** | **CUMULATIVE TOTALS** |
| 1 | $158,035 | $158,035 |
| 2 | $158,034 | $158,034 |
| 3 | $156,263 | $156,263 |
| 4 | $154,495 | $154,495 |
| 5 | $155,722 | $155,722 |

Recommended future year total cost support, subject to the availability of funds and satisfactory progress of the project

**Fiscal Information:**

**Payment System Identifier:** 1731328881A1
**Document Number:** KMH135213A
**PMS Account Type:** P (Subaccount)
**Fiscal Year:** 2024

| IC | CAN | 2024 | 2025 | 2026 | 2027 | 2028 |
|---|---|---|---|---|---|---|
| MH | 8472598 | $158,035 | $158,034 | $156,263 | $154,495 | $155,722 |

Recommended future year total cost support, subject to the availability of funds and satisfactory progress of the project

**NIH Administrative Data:**
**PCC**: AK-TN / **OC**: 41033 / **Released**: Mercogliano, Theresa 11/02/2023
**Award Processed:** 11/10/2023 12:01:49 AM

#### SECTION II – PAYMENT/HOTLINE INFORMATION – 1K23MH135213-01

For payment and HHS Office of Inspector General Hotline information, see the NIH Home Page at http://grants.nih.gov/grants/policy/awardconditions.htm

#### SECTION III – STANDARD TERMS AND CONDITIONS – 1K23MH135213-01

This award is based on the application submitted to, and as approved by, NIH on the above-titled project and is subject to the terms and conditions incorporated either directly or by reference in the following:

a. The grant program legislation and program regulation cited in this Notice of Award.

b. Conditions on activities and expenditure of funds in other statutory requirements, such as those included in appropriations acts.
c. 45 CFR Part 75.
d. National Policy Requirements and all other requirements described in the NIH Grants Policy Statement, including addenda in effect as of the beginning date of the budget period.
e. Federal Award Performance Goals: As required by the periodic report in the RPPR or in the final progress report when applicable.
f. This award notice, INCLUDING THE TERMS AND CONDITIONS CITED BELOW.

(See NIH Home Page at http://grants.nih.gov/grants/policy/awardconditions.htm for certain references cited above.)

**Research and Development (R&D):** All awards issued by the National Institutes of Health (NIH) meet the definition of "Research and Development" at 45 CFR Part§ 75.2**.** As such, auditees should identify NIH awards as part of the R&D cluster on the Schedule of Expenditures of Federal Awards (SEFA). The auditor should test NIH awards for compliance as instructed in Part V, Clusters of Programs. NIH recognizes that some awards may have another classification for purposes of indirect costs. The auditor is not required to report the disconnect (i.e., the award is classified as R&D for Federal Audit Requirement purposes but non-research for indirect cost rate purposes), unless the auditee is charging indirect costs at a rate other than the rate(s) specified in the award document(s).

An unobligated balance may be carried over into the next budget period without Grants Management Officer prior approval.

This grant is subject to Streamlined Noncompeting Award Procedures (SNAP).

This award is subject to the requirements of 2 CFR Part 25 for institutions to obtain a unique entity identifier (UEI) and maintain an active registration in the System for Award Management (SAM). Should a consortium/subaward be issued under this award, a UEI requirement must be included. See http://grants.nih.gov/grants/policy/awardconditions.htm for the full NIH award term implementing this requirement and other additional information.

This award has been assigned the Federal Award Identification Number (FAIN) K23MH135213. Recipients must document the assigned FAIN on each consortium/subaward issued under this award.

Based on the project period start date of this project, this award is likely subject to the Transparency Act subaward and executive compensation reporting requirement of 2 CFR Part 170. There are conditions that may exclude this award; see http://grants.nih.gov/grants/policy/awardconditions.htm for additional award applicability information.

In accordance with P.L. 110-161, compliance with the NIH Public Access Policy is now mandatory. For more information, see NOT-OD-08-033 and the Public Access website: http://publicaccess.nih.gov/.

This award provides support for one or more clinical trials. By law (Title VIII, Section 801 of Public Law 110-85), the "responsible party" must register "applicable clinical trials" on the ClinicalTrials.gov Protocol Registration System Information Website. NIH encourages registration of all trials whether required under the law or not. For more information, see http://grants.nih.gov/ClinicalTrials_fdaaa/

Recipients must administer the project in compliance with federal civil rights laws that prohibit discrimination on the basis of race, color, national origin, disability, age, and comply with applicable conscience protections. The recipient will comply with applicable laws that prohibit discrimination on the basis of sex, which includes discrimination on the basis of gender identity, sexual orientation, and pregnancy. Compliance with these laws requires taking reasonable steps to provide meaningful access to persons with limited English proficiency and providing programs that are accessible to and usable by persons with disabilities. The HHS Office for Civil Rights provides guidance on complying with civil rights

Version: 13 - 8/3/2022 12:59 PM | Generated on: 11/10/2023 12:01 AM laws enforced by HHS. See https://www.hhs.gov/civil-rights/for-providers/provider-obligations/index.html and https://www.hhs.gov/.

- Recipients of FFA must ensure that their programs are accessible to persons with limited English proficiency. For guidance on meeting the legal obligation to take reasonable steps to ensure meaningful access to programs or activities by limited English proficient individuals, see https://www.hhs.gov/civil-rights/for-individuals/special-topics/limited-english-proficiency/fact-sheet-guidance/index.html and https://www.lep.gov.
- For information on an institution's specific legal obligations for serving qualified individuals with disabilities, including providing program access, reasonable modifications, and to provide effective communication, see http://www.hhs.gov/ocr/civilrights/understanding/disability/index.html.
- HHS funded health and education programs must be administered in an environment free of sexual harassment; see https://www.hhs.gov/civil-rights/for-individuals/sex-discrimination/index.html. For information about NIH's commitment to supporting a safe and respectful work environment, who to contact with questions or concerns, and what NIH's expectations are for institutions and the individuals supported on NIH-funded awards, please see https://grants.nih.gov/grants/policy/harassment.htm.
- For guidance on administering programs in compliance with applicable federal religious nondiscrimination laws and applicable federal conscience protection and associated anti-discrimination laws, see https://www.hhs.gov/conscience/conscience-protections/index.html and https://www.hhs.gov/conscience/religious-freedom/index.html.

In accordance with the regulatory requirements provided at 45 CFR 75.113 and Appendix XII to 45 CFR Part 75, recipients that have currently active Federal grants, cooperative agreements, and procurement contracts with cumulative total value greater than $10,000,000 must report and maintain information in the System for Award Management (SAM) about civil, criminal, and administrative proceedings in connection with the award or performance of a Federal award that reached final disposition within the most recent five-year period. The recipient must also make semiannual disclosures regarding such proceedings. Proceedings information will be made publicly available in the designated integrity and performance system (currently the Federal Awardee Performance and Integrity Information System (FAPIIS)). Full reporting requirements and procedures are found in Appendix XII to 45 CFR Part 75. This term does not apply to NIH fellowships.

**Treatment of Program Income:**
Additional Costs

#### SECTION IV – MH SPECIFIC AWARD CONDITIONS – 1K23MH135213-01

Clinical Trial Indicator: Yes
This award supports one or more NIH-defined Clinical Trials. See the NIH Grants Policy Statement Section 1.2 for NIH definition of Clinical Trial.

**AWARD NOTICE:**
This award has been made in response to the application submitted under the Funding Opportunity Announcement PA-20-206 which can be referenced at: https://grants.nih.gov/grants/guide/pa-files/PA-20-206.html.

**DATA SHARING PLAN:**
This award is subject to the data sharing guidance outlined in NOT-MH-23-100 and can be found at: https://grants.nih.gov/grants/guide/notice-files/NOT-MH-23-100.html. The recipient will adhere to the resource and data sharing plan. Dissemination of study data will be in accord with the Recipient's accepted data sharing plan. **Please note that a statement of progress on the Sharing Plan must be included in the Research Performance Progress Report (RPPR) under section C.5 "Other Products and Resource**

**Sharing.”** Failure to adhere to the sharing plan as mutually agreed upon by the Recipient and the NIH/IC may result in Enforcement Actions as described in the NIH Grants Policy Statement.

Complete NIMH data sharing terms and conditions can be found at https://nda.nih.gov/contribute/sharing-regimen.html. Instructions are available at http://grants.nih.gov/grants/rppr/index.htm. Complete guidelines on data sharing are available at https://sharing.nih.gov/data-management-and-sharing-policy/about-data-management-and-sharing-policies/research-covered-under-the-data-management-sharing-policy#after.

**PARTICIPANT RECRUITMENT - MILESTONES:**
Future NIMH support for this study is contingent upon adequate participant recruitment based on projected milestones as approved in the Recruitment Milestone Reporting system (RMR) on 9/21/2023. It is expected that 8 of the 102 total projected participants will be recruited by 8/01/2024. This tri-yearly recruitment report should be submitted electronically to NIMH after each milestone period of April 1, August 1 and December 1 at: http://wwwapps.nimh.nih.gov/rmr/displayHome.action. In the event that actual recruitment falls significantly below projected milestones, NIMH may consider withholding future support and/or negotiating an orderly phase-out of this study. Information regarding the NIMH Policy for the Recruitment of Participants in Clinical Research is available at: https://grants.nih.gov/grants/guide/notice-files/NOT-MH-19-027.html.

**DATA AND SAFETY MONITORING:**
This grant is subject to the clinical research policies outlined in NOT-MH-19-027. The recipient will adhere to the NIH (NIH GPS 4.1.15) and NIMH policies (NOT-MH-19-027) including appropriately providing adequate data and safety monitoring and timely reporting of key events as defined in the NIMH Reportable Events Policy. The level and frequency of human subject data and safety monitoring should always be commensurate with the risk and nature of the research.

This grant requires an Independent Safety Monitor (ISM). The recipient and Principal Investigator(s) should review the NIMH Policy Governing Independent Safety Monitors and Independent Data and Safety Monitoring Boards (cited in NOT-MH-19-027) for details regarding roles, independence, responsibilities, and review reports. Each RPPR submitted must include, in Section **G.1 SPECIAL NOTICE OF AWARD TERMS AND FUNDING OPPORTUNITIES ANNOUNCEMENT REPORTING REQUIREMENTS**, copies of all ISM review determinations for all meetings/reviews conducted during the reporting period since the last annual award. The RPPR is expected to also include the following information: (1) the dates of the ISM review and name(s) of the ISM; (2) a summary of key safety indices (e.g., cumulative rates of AEs/SAEs) during the reporting period and cumulatively over the course of the project; and (3) a summary of any recommendations from the ISM and any actions taken based on the recommendations.

**GCP TRAINING:**
NIH expects that all NIH-funded investigators and staff who are involved in the conduct, oversight, or management of clinical trials should be trained in Good Clinical Practice (GCP), consistent with principles of the International Conference on Harmonization (ICH) as stated in NOT-OD-16-148.

**CLINICAL TRIAL DISSEMINATION PLAN:**
The clinical trial(s) supported by this award is subject to the plan within the competing application dated 2/13/2023 submitted to NIH and the NIH policy on Dissemination of NIH-Funded Clinical Trial Information. The plan states that the clinical trial(s) funded by this award will be registered in ClinicalTrials.gov not later than 21 calendar days after enrollment of the first participant and primary summary results reported in ClinicalTrials.gov, not later than one year after the completion date. The reporting of summary results is required by this term of award even if the primary completion date occurs after the period of performance.

**CLINICAL TRIAL REQUIREMENTS:**
This award is subject to additional certification requirements with each submission of the Annual, Interim, and Final Research Performance Progress Report (RPPR). The recipient must agree to the following annual certification when submitting each RPPR. By submitting the RPPR, the AOR signifies compliance, as follows:
In submitting this RPPR, the SO (or PD/PI with delegated authority), certifies to the best of his/her knowledge that, for all clinical trials funded under this NIH award, the recipient and all investigators conducting NIH-funded clinical trials are in compliance with the recipient's plan addressing compliance with the NIH Policy on Dissemination of NIH-Funded Clinical Trial Information. Any clinical trial funded in whole or in part under this award has been registered in ClinicalTrials.gov or will be registered not later than 21 calendar days after enrollment of the first participant. Summary results have been submitted to ClinicalTrials.gov or will be submitted not later than one year after the completion date, even if the completion date occurs after the period of performance.

**HUMAN SUBJECTS RESEARCH:**
This award includes human subject research studies and must conform to the DHHS policies for the Protection of Human Subjects research, which are a term and condition of award. Human subjects research is covered by the 2018 Common Rule, and may not be initiated until the associated protocols have received IRB approval as specified in 45 CFR 46. Failure to comply with the terms and conditions of award may result in the disallowance of costs and collected data and/or additional enforcement actions as outlined in Section 8.5 of the NIH Grants Policy Statement.

Data Management and Sharing Policy: Applicable

This project is expected to generate scientific data. Therefore, the Final NIH Policy for Data Management and Sharing applies. The approved Data Management and Sharing (DMS) Plan is hereby incorporated as a term and condition of award, and the recipient shall manage and disseminate scientific data in accordance with the approved plan. Any significant changes to the DMS Plan (e.g., new scientific direction, a different data repository, or a timeline revision) require NIH prior approval. Failure to comply with the approved DMS plan may result in suspension and/or termination of this award, withholding of support, audit disallowances, and/or other appropriate action. See NIH Grants Policy Statement Section 8.2.3 for more information on data management and sharing expectations.

| Budget | Year 1 | Year 2 | Year 3 | Year 4 | Year 5 |
|---|---|---|---|---|---|
| DMS Costs | $9,372 | $1,200 | $1,200 | $1,200 | $2,400 |

**SPREADSHEET SUMMARY**
**AWARD NUMBER:** 1K23MH135213-01

**INSTITUTION:** LAUREATE INSTITUTE FOR BRAIN RESEARCH

| Budget | Year 1 | Year 2 | Year 3 | Year 4 | Year 5 |
|---|---|---|---|---|---|
| Salaries and Wages | $77,063 | $77,063 | $77,063 | $77,063 | $77,063 |
| Fringe Benefits | $19,266 | $19,266 | $19,266 | $19,266 | $19,266 |
| Personnel Costs (Subtotal) | $96,329 | $96,329 | $96,329 | $96,329 | $96,329 |
| Other | $50,000 | $49,999 | $48,359 | $46,722 | $47,858 |
| TOTAL FEDERAL DC | $146,329 | $146,328 | $144,688 | $143,051 | $144,187 |
| TOTAL FEDERAL F&A | $11,706 | $11,706 | $11,575 | $11,444 | $11,535 |

| | | | | | |
|---|---|---|---|---|---|
| TOTAL COST | $158,035 | $158,034 | $156,263 | $154,495 | $155,722 |

| Facilities and Administrative Costs | Year 1 | Year 2 | Year 3 | Year 4 | Year 5 |
|---|---|---|---|---|---|
| F&A Cost Rate 1 | 8% | 8% | 8% | 8% | 8% |
| F&A Cost Base 1 | $146,329 | $146,328 | $144,688 | $143,051 | $144,187 |
| F&A Costs 1 | $11,706 | $11,706 | $11,575 | $11,444 | $11,535 |
