## Supplementary E for "Interoceptive mechanisms of body image disturbance in anorexia nervosa: Study protocol of a randomized parallel-group trial"

IRB APPROVED
AS MODIFIED
Sep 16, 2024

### RESEARCH PARTICIPANT INFORMATION AND CONSENT FORM

**TITLE:** Interoceptive mechanisms of body image disturbance in anorexia nervosa

**PROTOCOL NO.:** 2023-007
WCG IRB Protocol #20234215
2023-007

**SPONSOR:** Laureate Institute for Brain Research

**INVESTIGATOR:** Emily Choquette, PhD
6655 S Yale Ave
Tulsa OK 74136
USA

**STUDY-RELATED PHONE NUMBERS:** Emily Choquette, PhD
918-240-2583
918-481-4000 (24 hours)

Taking part in this research is voluntary. You may decide not to participate, or you may leave the study at any time. Your decision will not result in any penalty or loss of benefits to which you are otherwise entitled.

If you have any questions, concerns, or complaints or think this research has hurt you, talk to the research team at the phone number(s) listed in this document.

#### RESEARCH CONSENT SUMMARY

You are being asked for your consent to take part in a research study. This document provides a concise summary of this research. It describes the key information that we believe most people need to decide whether to take part in this research. Later sections of this document will provide all relevant details.

##### What should I know about this research?

- Someone will explain this research to you.
- Taking part in this research is voluntary. Whether you take part is up to you.
- If you don't take part, it won't be held against you.
- You can take part now and later drop out, and it won't be held against you
- If you don't understand, ask questions.
- Ask all the questions you want before you decide.

IRB APPROVED
AS MODIFIED
Sep 16, 2024

**How long will I be in this research?**
We expect that your taking part in this research will last 28-32 weeks.

**Why is this research being done?**
The purpose of this research is to study the effects of an intervention focused on helping individuals with anorexia nervosa interact with their body, thoughts, and emotions using acceptance and mindfulness based skills.

**What happens to me if I agree to take part in this research?**
If you decide to take part in this research study, the general procedures include attending intervention sessions in the clinic, answering questionnaires and interview questions, completing behavioral tasks, and having an MRI.

**Could being in this research hurt me?**
The most important risks or discomforts that you may expect from taking part in this research include feeling uncomfortable from the questions asked, worsening mental health symptoms or behaviors, risks related to salt water exposure including slipping and falling and mild skin irritation, having feelings of claustrophobia and anxiety from the MRI, and loss of confidentiality.

**Will being in this research benefit me?**
We cannot promise any benefit. Previous studies on similar interventions have indicated the intervention may result in reduced anxiety and disordered eating symptoms.

Possible benefits to others include learning more about potential of interventions focused on helping individuals interact differently with their thoughts, emotions, and body and how these may be helpful for treatment of certain eating disorders.

**What other choices do I have besides taking part in this research?**
Instead of being in this research, your choices may include other options for treatment for anorexia nervosa or other symptoms and conditions you may be experiencing including other talk therapy or medications.

#### DETAILED RESEARCH CONSENT

You are being invited to take part in a research study. A person who takes part in a research study is called a research subject, or research participant.

In this consent form “you” generally refers to the research subject. If you are being asked as the parent, or guardian to permit the subject to take part in the research, “you” in the rest of this form generally means the research subject.

**What should I know about this research?**

- Someone will explain this research to you.
- This form sums up that explanation.
- Taking part in this research is voluntary. Whether you take part is up to you.
- You can choose not to take part. There will be no penalty or loss of benefits to which you are otherwise entitled.
- You can agree to take part and later change your mind. There will be no penalty or loss of benefits to which you are otherwise entitled.
- If you don't understand, ask questions.
- Ask all the questions you want before you decide.

**Why is this research being done?**

The purpose of this research is to study the effects of an intervention focused on helping individuals become more accepting of their thoughts, emotions, and body using acceptance and mindfulness based interventions. In one condition, this will be paired with floatation via Reduced Environmental Stimulation Therapy [i.e., floatation-REST]) compared to a video guided skills practice in people with anorexia nervosa. While based on validated principles, combining these acceptance and mindfulness-based skills with floatation-REST or skills videos is considered experimental therapy.

About 102 subjects will take part in this research.

**How long will I be in this research?**

We expect that your taking part in this research will last approximately 28 – 32 weeks. There will be two baseline visits approximately 3 hours in length, after which you will engage in a 1 hour introduction session, and three 3- hour intervention sessions during which you will answer questions before and after engaging in your assigned condition. There will also be two 3 hour post intervention sessions and two remote follow-up visits (lasting approximately 1.5 hours each).

**You may stop participating at any time.**

**What happens to me if I agree to take part in this research?**

| |
|---|
| **Consent and Screening**<br>Consent<br>Authorization for medical record release<br>Medical and Demographics Information<br>Vital Signs |
| **Visits 1-2: Baseline***<br>Self-report questionnaires<br>Urine and Breathalyzer Screen<br>Vital Signs<br>Behavioral Tasks<br>Neuroimaging (fMRI) |
| **Visit 3: Intervention**<br>Introduction Session with trained therapist<br>Self-report questionnaire |
| **Visits 4 - 6: Intervention**<br>Pre Session Questionnaires<br>Either IFT or EFT sessions<br>Post Session Questionnaires |
| **Visits 7-8: Post Intervention***<br>Questionnaires<br>Behavioral Tasks<br>Feedback Interview<br>Neuroimaging (fMRI) |
| **Visits 9 & 10: 6 week and 6 month follow-up**<br>Questionnaires<br>Feedback Interview (Visit 10 only) |

*Please note that in some cases, baseline and post-intervention visits may be divided into multiple shorter sessions.

If you agree to take part in the study you will have the following tests and procedures at the Laureate Institute for Brain Research (LIBR):

***Interventions:*** You will receive either acceptance and mindfulness-based intervention paired with floatation-REST or practice video sessions. All participants will receive a one-hour in person acceptance and mindfulness based introduction session with a study therapist. During this session, you will be taught exercises to help you be more mindful of your current experience, specifically your thoughts, emotions, and bodily sensations. You have a 1 in 2 chance of being assigned to either treatment group. For both you will come to the LIBR Float Clinic and Research Center (FCRC) once or twice per week for 3 visits, each session will consist of approximately 15 minutes with a study therapist to review skills learned in the introduction

IRB APPROVED
AS MODIFIED
Sep 16, 2024

session and learn new skills. Then you will spend approximately 45 minutes in your assigned condition. Followed by a brief debrief (approximately 15 minutes) with the study therapist to process your experience.

*Floating:* You will be asked to lay in circular float pools which contain about 1 foot of water that is mixed with Epsom salts, allowing you to float on your back while lying in the water. You will be given the option to wear a swimsuit or be in the pool without clothes. Your session in the pool will be private. You will have your own private bathroom and shower for use during the float session.

While floating, both the light and sound will be reduced. You will be asked to practice the skills learned in the introduction session. The researcher will assist you with the preparations for the float and will be available to verbally communicate with you during the float session if you need assistance. Your privacy will be protected and maintained during the entire float experience. You will be asked to float for approximately 45 minutes one to two times per week for 3 visits.

*Video Practice:* You will be asked to view and practice along with skills videos related to reducing judgment of your thoughts and being more mindful of your current experience. You will be asked to practice each skill for a set period of time and respond to writing prompts about the exercise.

***Questionnaires:*** During the baseline, intervention, post intervention, and remote follow-up visits you will be asked to complete questionnaires. Depending on the visit questionnaires may take approximately 20 to 90 minutes to complete depending on the visit.

***Urine Screen:*** A urine drug and pregnancy screen will be completed at baseline to determine eligibility. You will not be eligible the study if either the pregnancy or drug screen are positive. A second urine pregnancy screen will be completed at post-intervention visit two prior to the MRI scanning.

***Physiological Measures:*** Your blood pressure and heart rate will be taken during several sessions and your heart rate will be monitored during one of the behavioral tasks in which you are asked to count your heart beat. Electrocardiogram (ECG) is a safe, non-invasive measure, of heart rate.

***Magnetic Resonance Imaging (MRI):*** For your safety, you will be given a brief pre-screening form to complete. It will tell us if you have any metal inside your body. The pre-screening form may identify some unanticipated medical findings that prevent you from participating in the MRI scan. If so, these findings will remain as confidential as possible, and will be discussed with you by a researcher involved in this study.

IRB APPROVED
AS MODIFIED
Sep 16, 2024

The MRI scanner rapidly takes pictures of your brain without exposure to harmful ionizing radiation. The MRI scanner is a metal cylinder surrounded by a strong magnetic field. During the MRI, you will lie on a table that can slide in and out of the cylinder. You will be asked to lie still during scanning by remaining as relaxed as possible. While in the scanner you will hear loud knocking noises, and you will be fitted with earplugs to muffle the sound. You will be able to communicate with the MRI staff at all times during your scan, and you may ask to be moved out of the machine at any time. We may also monitor your heart rate and respiration using a device on your finger and a rubber belt that fits loosely around your chest using the built-in scanner equipment.

The study will use anatomical MRI and functional Magnetic Resonance Imaging (fMRI). During the fMRI, the MRI scanner measures changes in blood oxygen levels in different brain regions while you perform mental tasks. The tasks involve attending to body sensations and viewing images and making ratings about them. Other tasks involve resting in the scanner without performing any activity. This procedure will provide us with information about which brain areas become more active (that is, receive more blood oxygen) while you are performing the tasks.

There will be MRI scanning sessions during Visits 2 and 8. The time in the scanner will be approximately 90 minutes per visit.

***Behavioral Tasks****:* You will be asked to complete several behavioral tasks related to perception of your environment and your bodily signals. Body measurements will be taken as part of the tasks. These may include counting your heartbeats, estimating body size, and holding your breath.

**What are my responsibilities if I take part in this research?**
If you take part in this research, you will be responsible to:

- Alert the study team if you believe you are or could become pregnant during the study
- Alert the study team if your medications change within one week of the prescribed intervention
- Alert the study team if you are currently taking recreational substances or medications not as prescribed.

**Could being in this research hurt me?**
There is risk of loss of confidentiality.

*Risks associated with Questionnaires and Screening:* Risk of questionnaires and screening evaluation are minimal and most likely risks include fatigue, discomfort, or irritation. Some of the questions may be distressing or uncomfortable to answer and may cause mild psychological distress. You have the right to refuse to answer any question.

IRB APPROVED
AS MODIFIED
Sep 16, 2024

*Behavioral Tasks*: During this study, body measurements will be taken. This may cause some emotional distress. To minimize discomfort body size and composition measurements will be taken from behind you and not shared with you or anyone else outside of study staff. Another behavioral task will require you to hold your breath which may induce an acute state of discomfort or lightheadedness which dissipates quickly after the task is complete. Finally, you will be asked to identify your heart beat during one of the behavioral tasks which may cause frustration if you have difficulty doing so.

*Floatation-REST:* There are no known long-term risks or consequences of floating. Minor risks that have been associated with floating may include:

- slipping and falling due to wet environment
- feeling disoriented or scared due to the dark
- occasional neck discomfort while floating
- brief visual or auditory hallucinations in a small subset of participants (which are typically reported as pleasant)
- minor ear ache related to the salt crystals drying in the ear canal
- stinging of wounds or cuts from the salt water
- burning in the eyes or a bitter taste in the mouth if directly exposed to the salt water
- mild skin irritation due to the salt
- feeling uncomfortable being naked
- general feelings of boredom

*Acceptance and mindfulness-based intervention:* Therapeutic interventions can cause initial discomfort or distress due to the sensitive nature of topics covered. It is possible that symptoms may not improve or will worsen during the course of the intervention.

*Physiological Measurements:* ECGs are safe, non-invasive measure. Electrodes are attached using sticky patches to the skin may leave a small indent or mild skin irritation on the your body that will dissipate once the device is removed. The electrode gel might leave a sticky residue that can be easily washed off with warm water.

*Recording:* Some of the behavioral tasks and intervention sessions will be recorded. There is a risk of loss of privacy and confidentiality associated with recording. All recordings will be stored securely on LIBR servers and only accessible to trained research staff.

*Neuroimaging using MRI:* People are at risk for injury from the MRI magnet if they have any of the following metal implants or fragments:

- pacemakers or other implanted electronic devices
- brain stimulators
- dental implants
- aneurysm clips (metal clips on the wall of a large artery)

IRB APPROVED
AS MODIFIED
Sep 16, 2024

- metallic prostheses (including metal pins and rods, heart valves, and cochlear implants)
- permanent eyeliner
- implanted delivery pump
- shrapnel fragments

You will be asked to complete an MRI screening form for the MRI scan. You will be screened for these implants or metal fragments before the study, and if you have any of them, you will not receive an MRI scan and cannot be in the study. Tell the study doctor if you are uncertain whether you have any metal objects in your body. In addition, all magnetic objects (for example, watches, coins, jewelry, and credit cards) must be removed before entering the MRI scanner room.

Welders and metal workers are also at risk for injury because they may be unaware of small metal fragments in the eye.

There are no known long-term risks or consequences of MRI scans. However, you may become uncomfortable because you will be lying in a small space. Some people are bothered by the loud thumping noises made by the scanner. You will wear earplugs to reduce the noise and increase your comfort during scanning. LIBR study staff will closely and continuously monitor you throughout the scanning procedure. You will be able to use an emergency call button at all times during the scan. You will be removed from the scanner immediately if you request to be removed.

If you are a female, it is best that you not be pregnant while participating in this study. A urine-based pregnancy test may be obtained prior to any MRI scanning.

However, all the risks of the study cannot be currently predicted.

#### Will being in this research benefit me?

There may or may not be any direct benefit to you from participating in this study. All subjects in the study will be receiving treatment and it is possible that you may experience a decrease in symptoms as a result of these procedures. However, benefit from the treatment cannot be guaranteed.

#### Will it cost me money to take part in this research?

Neither you not your health insurance will be charged for any of the study tests, procedures, or activities. If you need to be hospitalized, voluntary or involuntary, Laureate Institute for Brain Research does not intend to provide payment for this and you or your insurance provider will be billed for these costs.

IRB APPROVED
AS MODIFIED
Sep 16, 2024

#### Will I be paid for taking part in this research?

You will be paid $20 per hour of questionnaire, behavioral task, interview, and intervention sessions (or $10 per half hour). The estimated time across all study visits is about 22.5 hours. While you are prepped for the MRI scanner and in the MRI scanner you will be paid $50 per hour. Your time in prep and in the scanner is approximately 1.5 hours per scan.

You will be paid through a ClinCard (similar to a debit card). Payments will be deposited within 24-48 hours of the visit.

#### What other choices do I have besides taking part in this research?

This study is for research purposes only, so your other option is not to participate in the study.

While the therapies being provided as part of this study have been found to be beneficial for individuals experiencing anorexia nervosa to reduce body image concerns and anxiety, there are other options to obtain therapy for these symptoms or other symptoms and conditions you may be experiencing. Your options may include approved medications, as well as different talk therapies than used in the current study. Information concerning community clinics and treatment resources will be provided to you as requested.

#### What happens to the information collected for this research?

Your private information and your medical record will be shared with individuals and organizations that conduct or watch over this research, including:

- The research sponsor
- People who work with the research sponsor
- Government agencies, such as the Food and Drug Administration
- WCG IRB, the Institutional Review Board (IRB) that reviewed this research

We may publish the results of this research. However, we will keep your name and other identifying information confidential. We protect your information from disclosure to others to the extent required by law. We cannot promise complete secrecy.

A description of this clinical trial will be available on http://www.ClinicalTrials.gov, as required by U.S. Law. This Web site will not include information that can identify you. At most, the Web site will include a summary of the results. You can search this Web site at any time.

Data in this research might be deidentified and used for future research or distributed to another investigator for future research without your consent. Any data or results obtained from these future research will not be shared with you because it is not possible to predict how they will be utilized by other Investigators.

IRB APPROVED
AS MODIFIED
Sep 16, 2024

**National Institute Data Archive (NDA)**

- Data from this study will be submitted to the National Institute of Mental Health's data archive, a large database where deidentified study data is stored and managed.
- Deidentified study data means that all personal information (such as name, address, birthdate and phone number) will be removed and replaced with a code number.
- Once uploaded, other researchers can request your deidentified study data. Every researcher (and institutions to which they belong) who requests your deidentified study data must promise to keep your data safe and promise not to try to learn your identity. Experts at the NIH who know how to keep your data safe will review each request carefully to reduce risks to your privacy.
- Sharing your study data does have some risks, although these risks are rare. Your study data could be accidentally shared with an unauthorized person who may attempt to learn your identity. The study researchers will make every attempt to protect your identity.
- You may not benefit directly from allowing your study data to be shared with NDA. However, study data may help researchers around the world to better understand mental health.
- You may decide now or later that you do not want your study data to be added to the NDA. You can still participate in this research study even if you decide that you do not want your data to be added to the NDA. To opt out, please provide the study staff with a written letter requesting that your data not be submitted to the NDA database.
- Once your data is part of the NDA, the study researchers cannot take back the study data that was shared before they were notified that you changed your mind. If you would like more information about NDA, this is available on-line at http://nda.nih.gov.

**Certificate of Confidentiality**

We will do everything we can to keep others from learning about your participation in this study. To further help us protect your privacy, we have obtained a Certificate of Confidentiality from the United States Department of Health and Human Services (DHHS). With this Certificate, we cannot be forced (for example by court order or subpoena) to disclose information that may identify you in any federal, state, local, civil, criminal, legislative, administrative, or other proceedings. The researchers will use the Certificate to resist any demands for information that would identify you, except to prevent serious harm to you or others, and as explained below.

You should understand that a Certificate of Confidentiality does not prevent you, or a member of your family, from voluntarily releasing information about yourself, or your involvement in this study.

If an insurer or employer learns about your participation, and obtains your consent to receive research information, then we may not use the Certificate of Confidentiality to withhold this information. This means that you and your family must also actively protect your own privacy. Disclosure will be necessary, however, upon request of DHHS for the purpose of audit or evaluation, and is limited only to DHHS employees involved in the review.

Finally, you should know the research team can take steps, such as telling authorities (for example, the Police) if 1) you tell us of plans to really hurt yourself, 2) you tell us of plans to really hurt another person, or 3) we learn that a child or elder has been or is being abused or neglected.

#### Why will this information be used and/or given to others?

This information will be used and/or given to others to do the research, to study the results, and to make sure that the research was done correctly. If the results of this study are made public, information that identifies you will not be used.

#### What if I decide not to give permission to use and give out my health information?

Then you will not be able to be in this research study.

#### Can I be removed from this research without my approval?

The person in charge of this research can remove you from this research without your approval. Possible reasons for removal include:

- It is in your best interest
- You have a side effect that requires stopping the research
- You are or become pregnant
- You are currently using illicit substances
- The research is canceled by the FDA or the sponsor
- You are unable to keep your scheduled appointments

We will tell you about any new information that may affect your health, welfare, or choice to stay in this research.

#### What happens if I agree to be in this research, but I change my mind later?

Taking part in this study is voluntary. You may choose not to participate. Refusal to participate will involve no penalty or loss of benefits to which you are otherwise entitled. If you agree to participant then decide against it, you can withdraw for any reason and leave the study without penalty or loss of benefits entitled.

#### What if I am injured because of taking part in this research?

If you get hurt or sick from participating in this study, emergency medical treatment is available. In an emergency, call 911. Be sure to tell the emergency staff and other healthcare providers of your participation in this study. Contact Emily Choquette, PhD as soon as possible at 918-240-2583 or 918-481-4000 (24 hours) if you think you have a research-related injury or illness. You or your health insurance provider will be billed to cover the cost of the medical or emergency services provided. No funds have been set aside by the Laureate Institute for Brain Research to compensate you if you are hurt or get sick. However, you still have the right to bring a lawsuit if you think you were harmed and deserve compensation.

#### Who can answer my questions about this research?

Your contact person for the study is Emily Choquette, PhD. She can be reached during business hours at 918-240-2583 or.

If you have questions, about your participation in this study, concerns, or complaints about the study, or have a research-related injury, contact the study investigator, 918-240-2583 or 918-481-4000 (24 hours). For emergencies call 911.

If you have questions, concerns, or complaints, or think this research has hurt you or made you sick, talk to the research team at the phone number listed above on the first page.

This research is being overseen by WCG IRB. An IRB is a group of people who perform independent review of research studies. You may talk to them at 855-818-2289 or if:

- You have questions, concerns, or complaints that are not being answered by the research team.
- You are not getting answers from the research team.
- You cannot reach the research team.
- You want to talk to someone else about the research.
- You have questions about your rights as a research subject.

**I agree to deidentified data being shared with the NDA**

☐ Yes

☐ No

#### Statement of Consent:

I have read the information in this consent form. I have been given an opportunity to ask questions. All my questions about the study and my (my child's) participation in it have been answered.

I Agree To Participate (to allow my child to participate) In This Study

I authorize the use and disclosure of my (my child's) health information to the parties listed in the authorization section of this consent for the purposes described above.

By signing this consent form, I have not given up any of my/my child's legal rights.

IRB APPROVED
AS MODIFIED
Sep 16, 2024

**Consent and Assent Instructions:**
Consent: Subjects 18 years and older must sign on the subject line below

For subjects under 18, consent is provided by the parents or guardian. Written assent is required for subjects ages 13 through 17 years using the Assent section below.

- All children are required to assent
- If assent is obtained, have the person obtaining assent document assent on the consent form

Your signature documents your permission for you or your child named below to take part in this research.

__________________________________________
Subject Name (printed)

#### CONSENT SIGNATURE:

__________________________________________ __________________
Signature of Subject (18 years and older) Date

__________________________________________ __________________
Signature of Parent or Guardian (Subject under 18) Date

__________________________________________
Name of Parent or Guardian (Subject under 18)(printed)

__________________________________________ __________________
Signature of Person Conducting Informed Consent Discussion Date

__________________________________________
Name of Person Conducting Informed Consent Discussion (printed)

**ASSENT SECTION:**

**Statement of person conducting assent discussion:**

- I have explained all aspects of the research to the subject to the best of his or her ability to understand.
- I have answered all the questions of the subject relating to this research.
- The subject agrees to be in the research.
- I believe the subject's decision to enroll is voluntary.
- The study doctor and study staff agree to respect the subject's physical or emotional dissent at any time during this research when that dissent pertains to anything being done solely for the purpose of this research.

❑ I have explained the study to the extent compatible with the subject's capability, and the subject has agreed to be in the study.

________________________________________ ____________________

Signature of Person Conducting Assent Discussion — Date

________________________________________

Name of Person Conducting Assent Discussion (printed)

**Statement of Parent or Guardian:**

My child appears to understand the research to the best of his or her ability and has agreed to participate.

________________________________________ ____________________

Signature of Parent or Guardian — Date

**Assent Signature:**

I understand the research and agree to participate.

________________________________________ ____________________

Signature of Subject (13 - 17 years old) — Date
